# Spatiotemporal trends in self-reported mask-wearing behavior in the United States: Analysis of a large cross-sectional survey

**DOI:** 10.1101/2022.07.19.22277821

**Authors:** Juliana C Taube, Zachary Susswein, Shweta Bansal

## Abstract

**Background:** Face mask-wearing has been identified as an effective strategy to prevent transmission of SARS-CoV-2, yet mask mandates were never imposed nationally in the United States. This decision resulted in a patchwork of local policies and varying compliance potentially generating heterogeneities in the local trajectories of COVID-19 in the U.S. While numerous studies have investigated patterns and predictors of masking behavior nationally, most suffer from survey biases and none have been able to characterize mask-wearing at fine spatial scales across the U.S. through different phases of the pandemic.

**Objective:** Urgently needed is a debiased spatiotemporal characterization of mask-wearing behavior in the U.S. This information is critical to further assess the effectiveness of masking, evaluate drivers of transmission at different time points during the pandemic, and guide future public health decisions through, for example, forecasting disease surges.

**Methods:** We analyze spatiotemporal masking patterns in over eight million behavioral survey responses from across the United States starting in September 2020 through May 2021. We adjust for sample size and representation using binomial regression models and survey raking, respectively, to produce county-level monthly estimates of masking behavior. We additionally debias self-reported masking estimates using bias measures derived by comparing vaccination data from the same survey to official records at the county-level. Lastly, we evaluate whether individuals’ perceptions of their social environment can serve as a less biased form of behavioral surveillance than self-reported data.

**Results:** We find that county-level masking behavior is spatially heterogeneous along an urban-rural gradient, with mask-wearing peaking in winter 2021 and declining sharply through May 2021. Our results identify regions where targeted public health efforts could have been most effective and suggest that individuals’ frequency of mask-wearing may be influenced by national guidance and disease prevalence. We validate our bias-correction approach by comparing debiased self-reported mask-wearing estimates with community-reported estimates, after addressing issues of small sample size and representation. Self-reported behavior estimates are especially prone to social desirability and non-response biases and our findings demonstrate that these biases can be reduced if individuals are asked to report on community rather than self behaviors.

**Conclusions:** Our work highlights the importance of characterizing public health behaviors at fine spatiotemporal scales to capture heterogeneities that may drive outbreak trajectories. Our findings also emphasize the need for a standardized approach to incorporating behavioral big data into public health response efforts. Even large surveys are prone to bias; thus, we advocate for a social sensing approach to behavioral surveillance to enable more accurate estimates of health behaviors. Finally, we invite the public health and behavioral research communities to use our publicly available estimates to consider how bias-corrected behavioral estimates may improve our understanding of protective behaviors during crises and their impact on disease dynamics.

## Introduction

Human behavior plays a key role in infectious disease transmission [1, 2]. Individuals’ decisions to get vaccinated, reduce their contacts, or wear a face mask, for example, can have a tremendous impact on disease dynamics [3–5]. The COVID-19 pandemic has highlighted that we are grossly limited in our ability to accurately measure and predict human behavior in the face of a novel pathogen. Yet knowledge of how human behaviors vary over time and space is critical to assess the effectiveness of mitigation strategies, to forecast disease surges, and to parameterize coupled disease-behavior models [6]. In particular, there is a paucity of data on how the frequency of face mask-wearing varies across the U.S. over different phases of the pandemic. This lack of fine-scale spatiotemporal data has forced public health organizations to adopt an inefficient one-size-fits-all approach to encourage masking nationwide, rather than directing resources and messaging to areas with lowest uptake. Here, we identify spatiotemporal trends in self-reported data on mask-wearing behavior across the U.S. from a large survey distributed from September 2020 to May 2021.

Mask-wearing has been identified as an effective strategy to reduce transmission of SARS-CoV-2. At the individual level, masks decrease both the amount of viral particles dispersed by an infectious wearer and inhaled by an uninfected wearer [7]. Modeling studies at the population-level (e.g., [4, 8, 9]) have suggested that mask-wearing can limit SARS-CoV-2 transmission and COVID-19 deaths, including under scenarios where masks aren’t worn universally or aren’t completely effective at blocking transmission. Randomized controlled trials (e.g., [10]) have also demonstrated that mask-wearing is an effective community-level intervention against COVID-19. Despite limited information at the time, the Centers for Disease Control and Prevention (CDC) initially recommended mask-wearing on April 3, 2020 [11]. Lack of a national mandate, though, resulted in a heterogeneous landscape of mask policies across states, counties, towns, and even individual businesses in the U.S. [12, 13]. Compounded with this spatial heterogeneity in mandates is additional heterogeneity in compliance, documented by localized observational studies (e.g., [14]). Collection of systematic, accurate data on mask-wearing levels across the U.S. is therefore essential to informing our understanding of the role of mask-wearing in the U.S. COVID-19 pandemic trajectory.

To address this gap, researchers and organizations have implemented extensive surveys on human behavior, including mask-wearing (e.g., [15–17]). These surveys hold exciting promise, yet they have contributed relatively little to our understanding of human behavior due to significant sampling limitations. Larger surveys with sufficient power to detect trends at local geographic scales are often not designed to capture a representative sample of the population. Demographic biases arising from a non-representative sample can be addressed with standard statistical tools such as survey weights, but other forms of bias, particularly non-response and social desirability bias, are more challenging to correct. Surveys about salient public health issues are especially likely to suffer from response bias; COVID-19 cautious individuals may be overrepresented in a survey about COVID-19 behavior. However, without estimates of the proportion of individuals in a given region that are COVID-19 cautious, there is no way to use survey weights on this demographic. Likewise, respondents may be influenced by social desirability bias when self-reporting COVID-19 preventive behaviors such as vaccination, social distancing, or mask-wearing so that they respond in a manner deemed favorable by society despite being inaccurate [18]. Without observational or ground-truth data to validate survey responses, quantifying this social desirability bias is difficult. Furthermore, it is critical that ground-truth data to correct biases in health behavior be used at a fine spatial and temporal scale to avoid further exacerbation of biases (e.g., [19]).

The value of surveys on public health behaviors can be further restricted when data collection is at the national or state level. Coarse-grained spatiotemporal information on human behavior is of limited utility, providing only sparse insight into local trends. Collecting responses at the national or state level ignores spatial heterogeneity at these finer scales, preventing the identification of these local effects that can drive disease dynamics. Spatial heterogeneity in not only drivers of disease transmission like human behavior, but also disease prevalence, has been well-documented across pathogens (e.g., [20–22]). For example, differences in connectivity between counties or states can affect the timing and geographic scale of disease spread, while national scale mobility data elides these key patterns [23, 24]. Likewise, aggregation of vaccination data to the state level can hide spatial clustering of unvaccinated individuals, which undermines herd immunity and could drive sustained measles outbreaks in the U.S. [25, 26]. Despite the importance of detailed local estimates on drivers of disease incidence, few studies have analyzed human behavior during the COVID-19 pandemic nationally at these fine spatial scales. Furthermore, most surveys are not conducted for long enough to capture human behavior changes over time, leaving scant opportunity to assess the effects of changing public health messaging/guidance or disease prevalence on human behavior.

Here, we systematically characterize mask-wearing across the United States at a fine spatiotemporal scale for nine months using a national survey and account for the bias in this survey. By comparing survey demographics and vaccination statuses with accurate ground-truth data, we estimate and account for survey and response biases in our analysis of masking behavior. With these bias-corrected estimates, we characterize the spatiotemporal heterogeneity in masking behavior at the county-month level across the U.S. Finally, we examine the differences between self-reported and community-reported estimates of masking using an additional survey question, seeking to understand whether these two measures are good predictors of one another. Our results are the most precise estimates of masking in the United States during the COVID-19 pandemic, providing insight into local variation in behavior in response to public health messaging and changes in COVID-19 incidence.

## Methods

In this study, we seek to characterize the spatiotemporal heterogeneity in self-reported masking behavior in the United States from the fall of 2020 to the spring of 2021. Due to low sample size in some counties, we use Bayesian binomial regression models to estimate mask-wearing proportions each month. Recognizing that surveys are subject to several types of bias, we use raking and resampling of responses to correct for unrepresentative samples and self-reported vaccination status compared to ground-truth vaccination data to quantify non-response and social desirability biases. With these estimates, we are able to identify spatiotemporal trends in bias-corrected masking behavior and compare these values to reported community levels of masking in a different survey question.

### Survey data & processing

We analyzed self-reported mask-wearing survey responses for all 50 U.S. states and the District of Columbia using data from the U.S. COVID-19 Trends and Impact Survey (CTIS) [27]. The CTIS was created by the Delphi Research Group at Carnegie Mellon University and distributed through a partnership with Facebook. Beginning in September 2020, a random state-stratified sample of active Facebook users were invited daily to take the survey about COVID-19 and report how often they wore a mask in the past 5-7 days (the number of days changed from 5 to 7 on February 8, 2021). Answer options were (1) All, (2) Most, (3) Some, (4) A little, or (5) None of the time, or (6) I have not been in public in the last 5-7 days (Figure S13). To dichotomize these responses for an analysis of the proportion of respondents wearing masks, we dropped respondents who hadn’t been in public recently or did not respond to the masking question, and considered responses of “all” and “most of the time” as masking, and all other responses as not masking. This cutoff is reasonable considering the raw proportions of responses in each category for September thru May (Figure S14). Due to sample size constraints, we aggregated these responses to the county-month scale. We ignore potential heterogeneity at smaller temporal (weekly) and spatial (zipcode) scales due to limited sample size. By dichotomizing masking responses, we also lose information about the frequency with which people mask, though we do expect the effect of this choice to be minimal (see Supplement for details).

### Bayesian binomial regression model

Due to small sample sizes in some U.S. counties, we used Bayesian binomial regression models to develop reliable estimates of the proportion of individuals masking in a given county-month. Population density was used as a fixed effect; masking behavior has previously been linked to population density and this variable was easily available at the county scale [14, 28]. We fit separate models for each month, allowing for a temporal trend without explicitly modeling it by specifying a parametric form. We define *M*_*i*_ as the number of respondents masking in county *i* (e.g., respondents that masked most or all of the time in the past 5-7 days), *N*_*i*_ as the total number of respondents in county *i* (*M*_*i*_ ≤ *N*_*i*_), and *p*_*i*_ as the county-level probability of a response consistent with masking. We use the following model to estimate 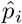 and 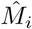:

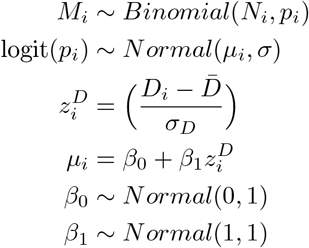

where *D*_*i*_ = log_10_(`population density`_*i*_) for county *i*. We ran the model using `brms` [29] with the `cmdstanR` 30 backend. We ran the sampler with 4 chains for 3000 iterations per chain. Sampler diagnostics indicate efficient exploration and that the model has converged: the n_eff_ *>* 950, n_eff_ per iteration ≥ 0.25, 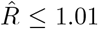, E-BFMI *>* 0.25, and no transitions hit max tree depth. All PSIS-LOO k statistic values are below 0.71, indicating that the model is robust to the influence of individual observations, and the distribution of Pareto *k* statistics is uniform, indicating that the model captures essential features of the data [31]. Posterior predictive checks indicate good model fit (Figure S15) as do plots of observed versus predicted and residual values (Figures S16, S17). We note that our binomial regression approach compensates for small sample sizes in some county-months, but the resulting estimates depend on the validity of our model structure.

We also explored more complex model specifications that included state or county-level random effects. However, both models suffered from lack of convergence or overfitting, and produced functionally similar results. Thus, we opted for the more parsimonious model presented above for our main findings; details on these additional models can be found in the supplement (Figures S20, S21).

### Survey raking and resampling

We were unable to use the provided weights for responses to the CTIS due to spatial and temporal mismatch with the scales of our data analysis. Thus, we calculated county-month weights for each observation using the `anesrake` package [32] and the U.S. Census American Community Survey’s 2018 data on county age, sex, and education distributions. Age, sex, and education distributions were based on each county’s population over the age of 18, to match the survey sample. We did not use race or ethnicity data in the raking scheme as its inclusion substantially reduced algorithm convergence but note that race/ethnicity was moderately correlated with education (Cramer’s V *>* 0.10). We then resampled from these responses using the calculated weights to estimate a raked masking proportion, which was fed into the binomial models, as described above. We excluded observations with missing age, sex, or education responses from the raking process and assigned equal weights to observations from county-months that did not converge (additional details in the Supplement).

### Estimation of CTIS masking bias

Given the likelihood of sampling, non-response, and social desirability biases, we generated bias-corrected estimates of masking in the U.S. In the absence of ground-truth masking data with which to calibrate these CTIS responses, we turned to a different survey question for which ground-truth data were available.

Beginning in late December 2020, the CTIS asked respondents whether they had received a COVID-19 vaccine. Response options were (1) Yes, (2) No, or (3) I don’t know. Meanwhile, ground-truth vaccination data were collected by combining state-reported and CDC data to estimate the percentage of people vaccinated in each county in the U.S. [33,34]. A comparison of CTIS responses and ground-truth vaccination data revealed that estimates of COVID-19 vaccination based on CTIS responses were much higher than true vaccination rates at the US county scale (Figures S18, S19, [19, 35]). Assuming that masking survey responses suffer from the same bias issues (in magnitude and direction) as vaccination responses, this result would suggest that CTIS responses also substantially overestimate masking behavior. Thus, we approximated survey bias by comparing the CTIS vaccination responses to the ground-truth vaccination data at the county-level and incorporating this bias into the model of CTIS masking behavior.

Like the masking data, the CTIS vaccination response data suffers from small and unrepresentative samples in some counties. Thus, we resampled the responses from April and May 2021 according to the survey weights we generated above and then used a (frequentist) binomial generalized linear mixed-effects model to estimate *p*_*i*_, the proportion of respondents who were vaccinated (assumed to be partial vaccination, with 1 of a 1-dose or 2-dose vaccine) at the county-level each week (details in Supplement).

Given these modeled CTIS county-level vaccination proportions, we compared them with the true vaccination data to calculate the expected bias in reported survey responses relative to ground-truth data in county *i*:

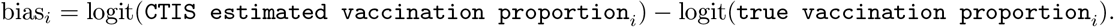

To increase the stability of our bias estimates, we used a linear mixed-effect model. This mixed effects model used random intercepts, which penalizes extreme coefficient estimates to the overall mean, and assumed that the residual error in the estimates was normally distributed. This model generates a penalized estimate of survey bias for each county from the difference in modeled reported vaccination and ground-truth vaccination:

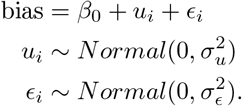

This model was implemented using `lmer` in the `lme4` package [36]. If there were no responses in the county *i* or a bias estimate could not be calculated, bias estimates for this county were imputed by taking the mean of surrounding county estimates.

We then incorporated these estimates into a Bayesian binomial regression model with an offset for bias to estimate the bias-corrected probability of reporting masking in county *i*. We define *M*_*i*_ as the number of respondents masking in county *i* out of *N*_*i*_ total respondents, and *p*_*i*_ as the county-level probability of a response consistent with masking. We use the following model to estimate 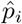 and 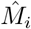:

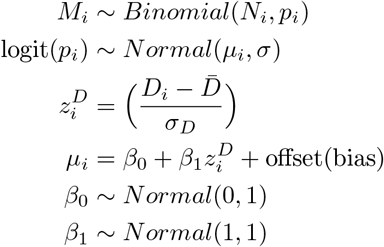

where *D*_*i*_ = log_10_(`population density`_*i*_) for county *i*. The bias-corrected proportion of individuals masking, *c*_*i*_, was calculated as

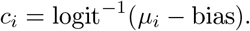

We ran the model using `brms` [29] with the `cmdstanR` [30] backend. We ran the sampler with 4 chains for 3000 iterations per chain. Sampler diagnostics indicate efficient exploration and that the model has converged: the n_eff_ *>* 1000, n_eff_ per iteration ≥ 0.3, and 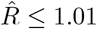.

### Community-reported masking

Beginning November 24, 2020, the CTIS asked a question about masking in one’s community: “In the past 7 days, when out in public places where social distancing is not possible, about how many people would you estimate wore masks?” The answer options were (1) All, (2) Most, (3) Some, (4) A few, or (5) None of the people, or (6) I have not been out in public places in the past 7 days. We dichotomized these responses and aggregated them to the county-month the same way as the self-reported CTIS masking responses for December 2020 through May 2021. We then modeled these community masking estimates the same way we modeled the CTIS masking data using Bayesian binomial regression and resampling weighted by survey weights but without a bias offset.

### Spatiotemporal analysis

All analyses were completed in R version 4.1.3 and maps were produced using choroplethr [37]. Urban-rural classes are from the National Center for Health Statistics’ Urban-Rural Classification Scheme [38].

### Ethical considerations

This study was reviewed by the Institutional Review Board at Georgetown University and was determined not to be human subjects research.

## Results

To characterize trends in masking behavior in the United States during the COVID-19 pandemic we used data from the COVID-19 Trends and Impact Survey (CTIS) conducted via Facebook from September 2020 through May 2021. Respondents self-reported how often they had worn a mask while in public in the last week (8,338,877 valid responses). We transformed these responses into a binary variable of masking or not masking and aggregated the responses to the county-month level to analyze spatiotemporal trends. To validate this data source, we analyzed a separate dataset from Outbreaks Near Me and found consistent spatiotemporal patterns (Figures S1, S2, S3), though both data sources suffer from issues of bias and low sample size. We addressed these issues in the CTIS data using binomial regression models to inform estimates of masking in counties with low sample size, and raking and sample rebalancing on age, gender, and education to adjust for unrepresentative samples. Recognizing that CTIS responses to a question about vaccination overestimated true vaccination rates, we quantified this bias for each county and used it to correct estimates of masking behavior, assuming that vaccination and masking behavior responses were equally as biased. (Vaccination and mask-wearing are both prosocial public health behaviors which are socially desirable to report and are likely correlated [39–43].) We analyzed overall spatial and temporal trends as well as fine-scale heterogeneity in the bias-corrected masking behavior estimates. Finally, we validated the bias-corrected CTIS values by comparing them to respondents’ estimates of the proportion of people masking in their community.

### Spatially heterogeneous effects of binomial regression model, survey raking, and debiasing

To demonstrate the spatially heterogeneous effects of our data processing scheme, Figure 1 shows the difference between estimates from three separate models and the raw CTIS masking data. We refer to this difference as the residual, though it is only an indicator of model fit in Figure 1A. In Figure 1B and C the residual values indicate where data corrections have caused the largest changes in estimates compared to the original data. After modeling the data with binomial regression, estimates of masking proportions are higher than observed values (Figure S4) in the central U.S., and slightly lower than observed values in the Northeast, Northwest, and Southwest (Figure 1A). Adjusting for unrepresentative samples with raking and resampling and rerunning the binomial regression model has a minor effect on mask-wearing estimates, only exhibiting a slight decrease compared to the model without raking (Figure 1B). Correcting for survey biases using vaccination data in the binomial regression model run on raked survey responses systematically decreases masking proportions, as expected and denoted by increased residuals (Figure 1C). (A map showing the spatial distribution of these biases is shown in Figure S19.) We refer to estimates from the model in Figure 1C as debiased or bias-corrected for the remainder of the paper. Our results reinforce that behavioral surveillance should be conducted carefully to limit bias initially.

**Figure 1.**
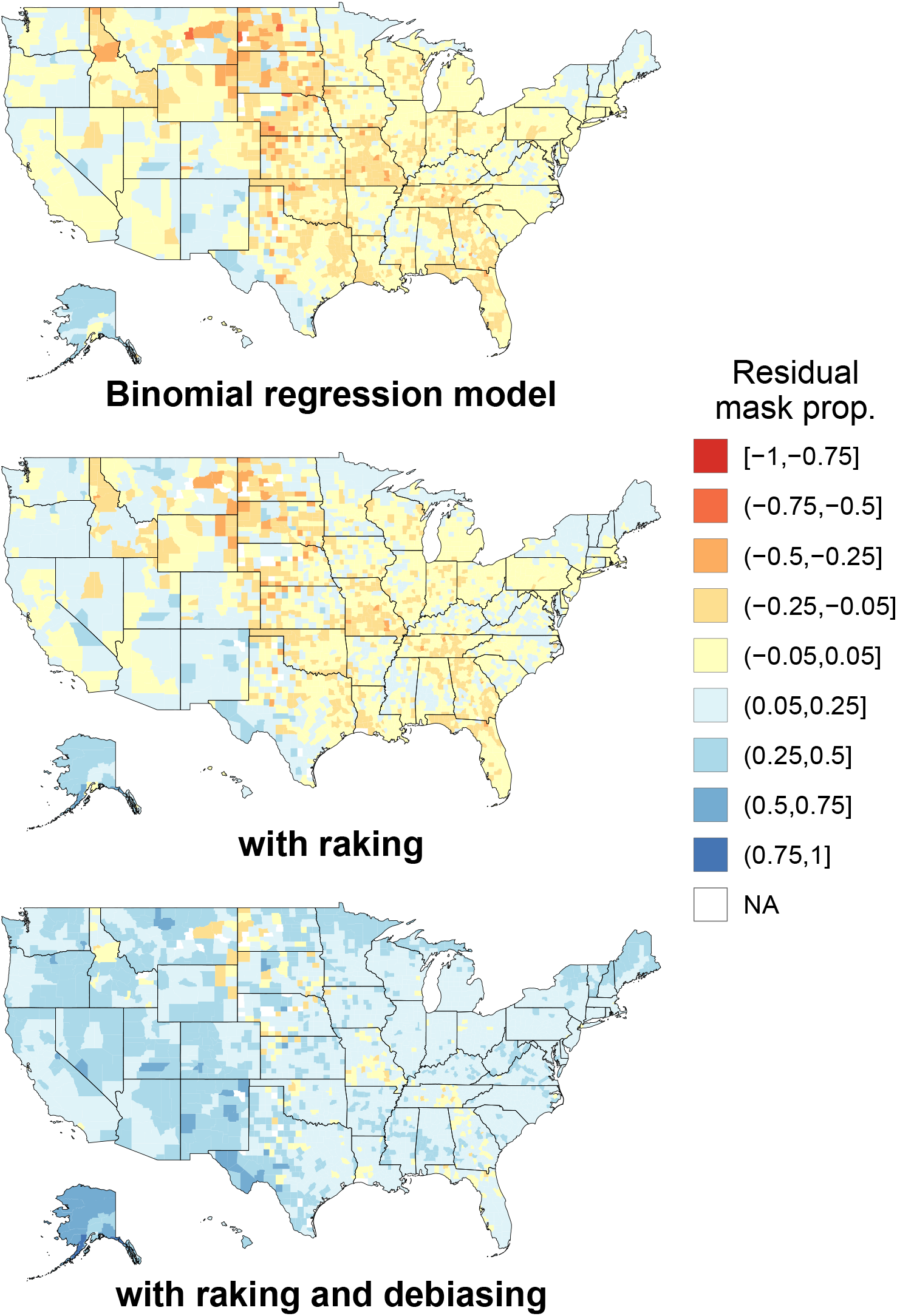
Visualization of spatially heterogeneous data processing effects. **(A)** Residuals following binomial regression model. **(B)** Residuals following binomial regression model with raking/sample rebalancing. **(C)** Residuals following binomial regression model with raking/sample rebalancing, and an offset for bias. Residuals are defined as the difference between the modeled and observed masking estimates at each analysis stage, where negative values indicate model estimates were higher than observed values, and positive residuals indicate model estimates were lower than observed values. All maps shown for February 2021.

### Masking behavior exhibits spatial and temporal heterogeneity and is positively associated with population density

Using bias-corrected masking proportions from the CTIS, we find that masking behavior is spatially heterogeneous over all months (Moran’s I between 0.68 and 0.70 for all months, Figures 2A, S5). Bias-corrected masking proportions range from 0.11 to 0.96, and vary substantially within states, emphasizing the importance of analyzing masking behavior at finer scales than the state or HHS region level. Masking proportions are closely linked to population density over the survey period: urban counties tend to have higher masking proportions than rural counties (Figure 2B). While masking proportions range quite a bit within NCHS urban-rural classifications, all differences between NCHS classes are significant (Kruskal-Wallis and Pairwise Wilcox test, n = 27,842, all *P* values<.001). Over all counties and survey months, the median fitted masking proportion in urban counties exceeds 0.8 while the median fitted masking proportion in the most rural counties is below 0.6.

**Figure 2.**
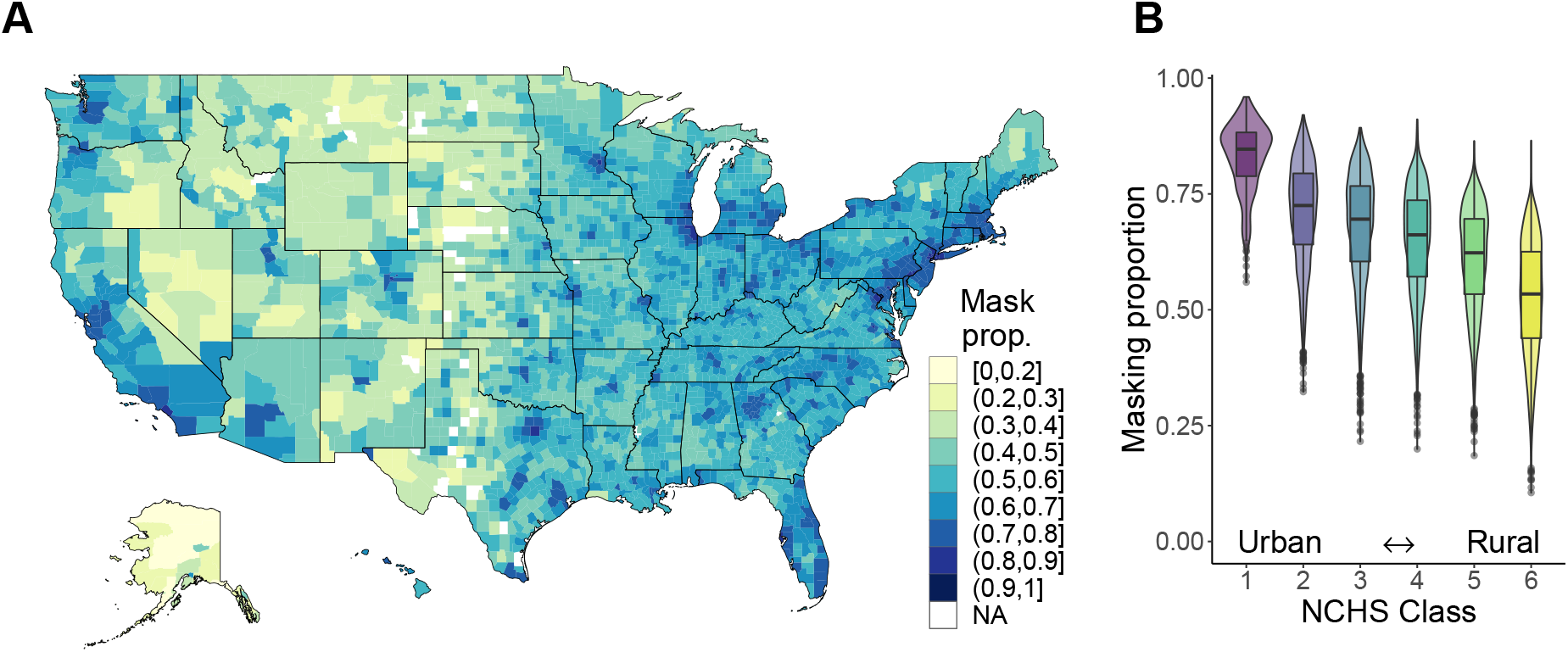
Bias-corrected masking behavior is spatially heterogeneous and higher in urban areas. **(A)** Map of bias-corrected masking behavior in October 2020 reveals high spatial heterogeneity. Masking proportions vary substantially even within a single state. Spatial heterogeneity does not notably vary over time (Figure S5). A selection of other months in the study period are shown in Figures S6, S7, and S8. **(B)** Breakdown of county masking proportions over all survey months by NCHS urban-rural classification. A direct relationship between median masking proportion and population density is observed.

Masking behavior not only varies geographically but also temporally. Peak masking behavior is observed in January 2021 while the lowest masking proportions are observed in May 2021 (Figure 3). Counties with higher mean masking proportions fluctuate less than counties with lower mean masking proportions from September to April but experience the largest differences from their mean values in May 2021. For context, we highlight that this decrease coincides with increasing proportions of vaccinated individuals in the U.S. (Figure 3, [33]), declining new infections [44], and decreasing reported worry about severe illness due to COVID-19 from the CTIS [27]. The policy context during this time was also shifting: On April 27, 2021 the CDC announced that fully vaccinated individuals no longer needed to wear masks outdoors [45] and on May 13, 2021 they announced that fully vaccinated individuals no longer had to wear masks indoors either [46]. Meanwhile, 49% of counties that ever had a mask mandate lifted it before May 1, 2021 (Figure S9). These announcements coincide with the observed decrease in masking in these months. Together, these analyses underscore the importance of tracking and analyzing mask wearing at fine spatial and across long temporal scales: further spatial and temporal aggregation of these data would have missed key heterogeneity previously not quantified and prevented future work from investigating the connection between policy and behavior change at appropriate granularity.

**Figure 3.**
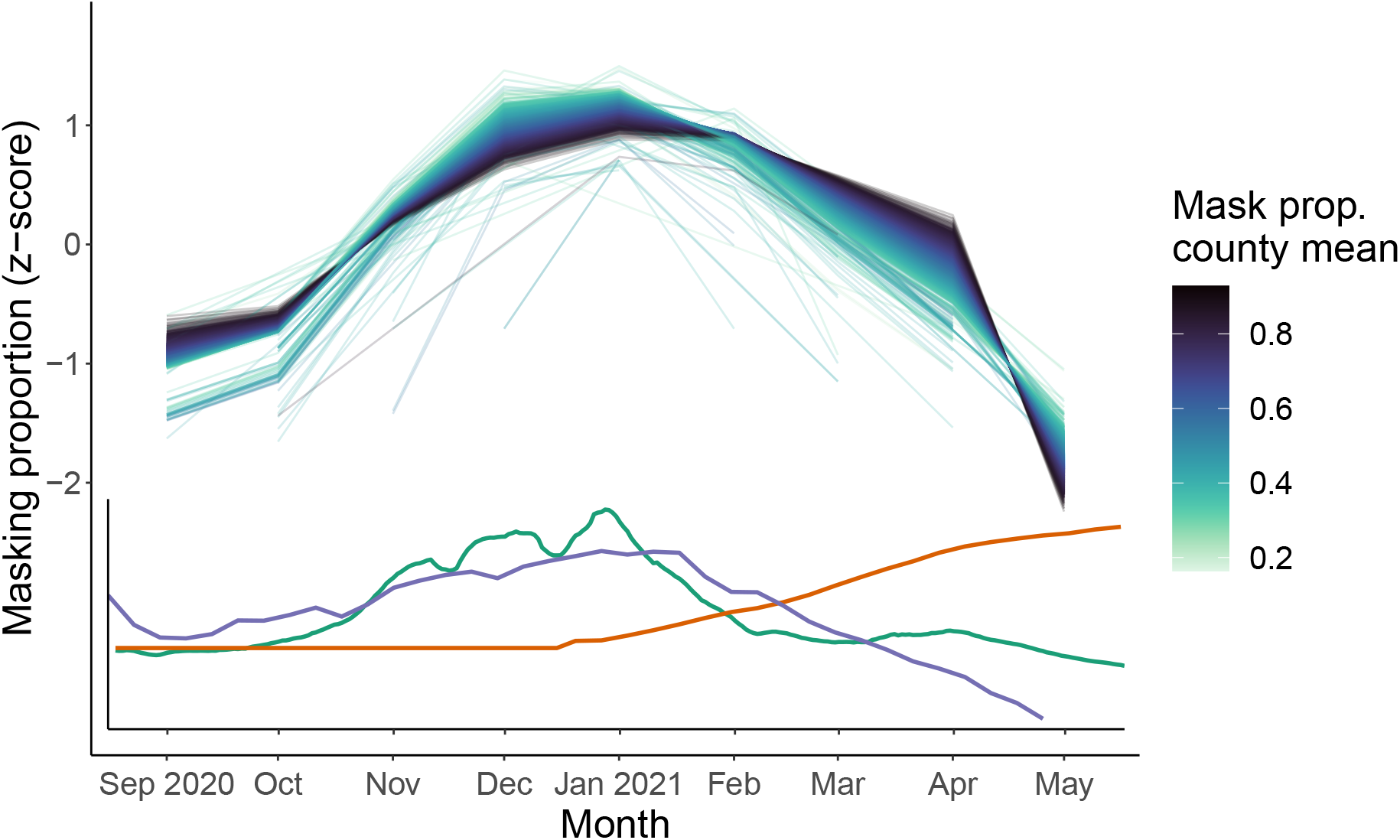
Bias-corrected masking behavior peaks in the winter of 2020-21 and falls in the spring of 2021, mirroring new cases and increasing vaccinations. Top curves show time series of z-score of bias-corrected masking proportions for each county colored by average masking proportion across the survey period. Inset plot shows z-scores of 7-day rolling average of new cases (green), proportion of individuals vaccinated nationally (orange), and reported worry about severe illness from COVID-19 in CTIS respondents (purple). Masking z-scores are based on the mean and standard deviation of each county’s masking estimates over the survey period.

### Community-reported masking levels are a good predictor of bias-corrected self-reported estimates

Bias-corrected masking proportions are well-approximated by modeled estimates of community-reported masking (Figure 4). The difference between the two mask-wearing proportion estimates ranges from − 6% to 5% and becomes more apparent in April and May 2021, particularly in rural areas. In May 2021, though, community estimates in urban areas tend to overestimate bias-corrected individual masking estimates. This result is quantitatively affected by influential observations but is qualitatively robust (Figure S10). These results suggest that surveying participants about community masking may give less biased responses than asking individuals to report their own masking behavior, potentially reducing social desirability bias and capturing parts of the population that may be otherwise less likely to respond to the survey.

**Figure 4.**
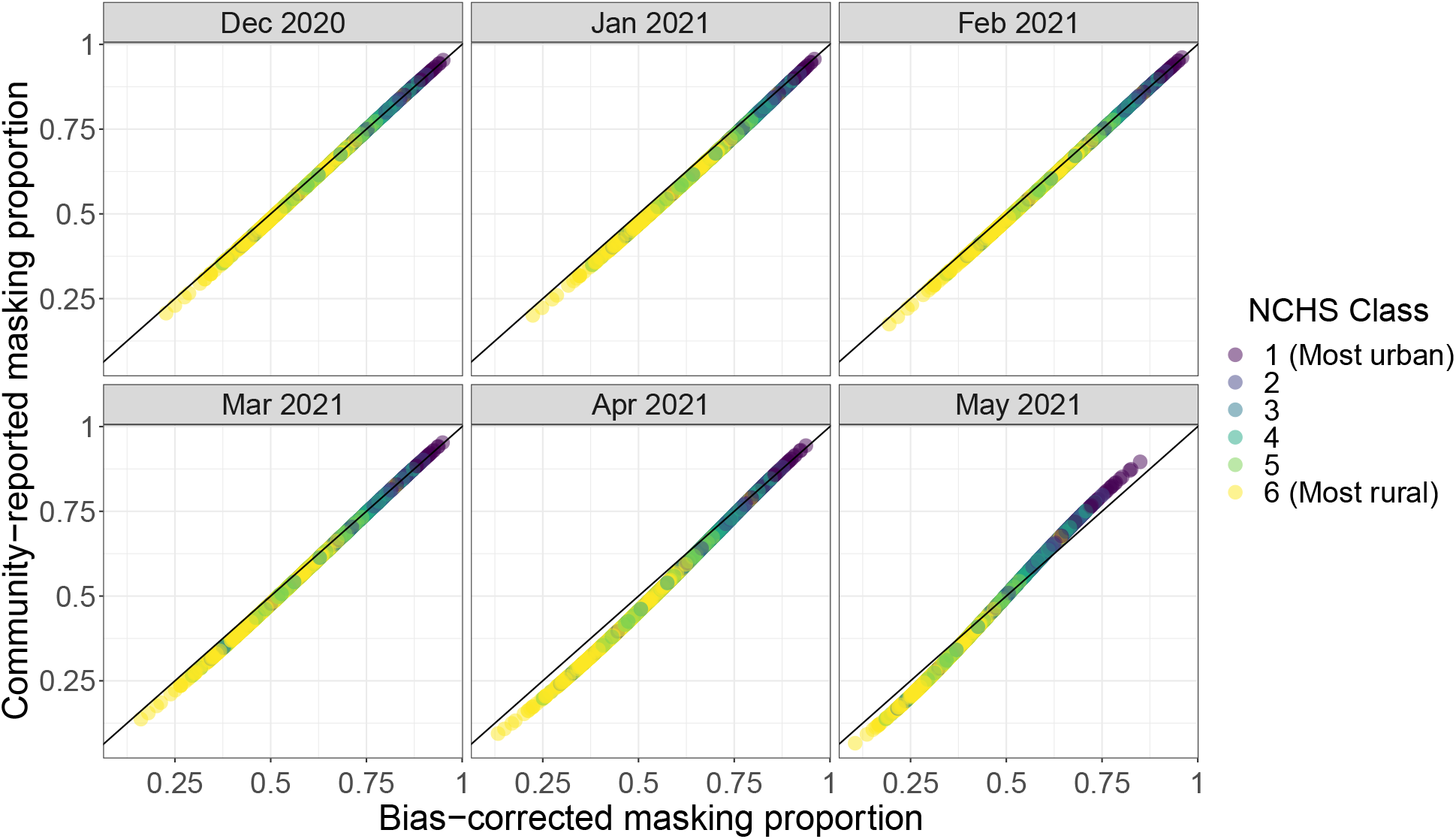
Community-reported masking gives a good estimate of bias-corrected self-reported masking. Community-reported masking refers to the CTIS question where individuals report how many people in their community are masking, which may decrease non-response and social desirability bias compared to asking individuals to self-report their masking behavior. Point color denotes urban-rural classes. Comparisons of individual and community-reported estimates at different analysis stages are shown in Figures S11, and S12.

## Discussion

Despite the widespread adoption of face mask-wearing at points during the COVID-19 pandemic in the U.S., the true prevalence of this behavior across temporal and spatial scales is largely unknown. Data on mask wearing has been collected through surveys, at varying spatiotemporal resolutions and with potentially varying survey biases (see e.g., [15–17]). Here, we characterize mask wearing behavior across the U.S. using self-reported masking data from a large national online survey. We employ Bayesian binomial regression models to remediate issues of small sample size, perform raking/sample balancing to address unrepresentative survey samples, and correct for additional response biases using measurable bias in vaccination data. We observe substantial spatial heterogeneity in masking behavior across urban versus rural counties with some temporal changes in mean masking estimates at the county-month level, most notably a steep decline in masking in May 2021. We find that community-reported masking responses well approximate our bias-corrected masking estimates. Other work adds to this validation: similar spatial heterogeneity is found in two other surveys, with some overlapping time periods (Figures S1, S2, S3, [17, 47]) and our debiased estimates generally agree with those recorded in observational studies, including higher levels of masking in urban areas (Table S1, [14, 28]). Our results reveal the landscape of masking behavior across three distinct phases of the pandemic (pre-surge, during the winter 2020-21 surge, and post-surge during the initial COVID-19 vaccination rollout). Our work also highlights the critical role that behavioral big data can play in pandemic response, if such data are used with caution.

Our findings have several implications for the fields of infectious disease epidemiology and public health policy. We identify high spatial and moderate temporal heterogeneity in masking behavior at the county-level – patterns that are obscured if data are aggregated to the state or HHS region-level. Contrary to our expectations, this level of spatial variability around the mean is consistent over time. Consequently, disease models should account for spatial variability in masking behavior but may only need to consider changes in masking dynamics over longer temporal scales. The high spatial heterogeneity we find in behavior also highlights the need for diverse and targeted public health approaches across the country rather than a single national program. Guidance set at the state-level without regard for differences in local conditions may miss early opportunities to control disease spread, prematurely enforce public health restrictions, and contribute to fatigue with public health restrictions. Thus we advocate for local behavioral data collection and geographically-targeted public health policy for optimized resource use and efficient disease suppression.

While county-level mask-wearing behavior varies across months, we observe little heterogeneity across counties in these temporal trends. The observed changes in masking behavior roughly correspond to national trends in new cases in the U.S. and self-reported worry about severe disease as reported in the CTIS, though we did not determine causality or examine this relationship at the individual or county level. Because we model county-level averages, this observed correlation could be driven by a specific demographic group or subset of individuals modifying their masking behavior rather than a uniform change in average mask uptake in a county’s population. The sharp decrease in masking in May 2021 is contemporaneous with many states lifting mask mandates (Figure S9) and an announcement from the CDC that vaccinated individuals no longer had to wear masks outdoors (April 27, 2021 [45]) or indoors (May 13, 2021 [46]). It is plausible that these policy changes could have impacted masking behavior, both in vaccinated and unvaccinated individuals, even though the change in CDC guidance did not apply to unvaccinated individuals [42, 48]. More work is needed to explore the potential differences in mask-wearing between vaccinated and unvaccinated individuals. Additional research could also focus on quantifying the impact of social norms on individuals’ masking behavior at fine spatiotemporal resolution in the U.S.

Recent work has highlighted the potential for big data sources to provide measurement of spatially disaggregated social phenomena (e.g., [49]), while other research points to the challenges of inferring high-quality estimates of behavior from such high-volume data sources [19]. In our work, we seek to steer away from “big data hubris” [50] by applying rigorous statistical methods to manage concerns about representativeness and bias, and by conducting an internal validation of our model-based estimates [51]. In particular, we address representativeness by age, sex, and education to capture socio-demographic response bias. Motivated by an association between COVID-19 preventative behaviors [39], we debias our masking estimates based on vaccination data to address additional non-response bias and social desirability bias. Finally, we internally validate our population-scale masking estimates of self-reported behaviors with responses of community behaviors, which may be subject to less non-response and social desirability bias than self-reporting questions [52]. We find that community-reported masking estimates agree closely with bias-corrected self-reported masking behavior, highlighting that surveying participants about community behavior may be an avenue to reduce survey bias. We note, however, that this finding may not apply in all settings; self-reported masking behavior on a university campus closely matched observed masking levels and questions about community masking were less accurate [53]. While these implications for analysis of surveys on human behavior may not apply universally, similar results have been found in other infectious disease applications including disease surveillance (using the CTIS data [27, 54]) and early outbreak detection in social networks [55, 56]. Our results further emphasize the promise of human social sensing going forward to make big data sources more meaningful [57].

Nevertheless, our approach has some limitations that are important to consider. We were unable to deal with all representation or response biases, including the exclusion of individuals under 18 years of age, lack of representativeness due to factors other than age, sex and education, recall bias, dishonest responses, and other characteristics that may be predictive of non-response or social desirability bias, such as political leanings or belief in COVID-19 conspiracies [27]. Likewise, we could not account for how individuals with Facebook accounts may engage differently in COVID-19 preventive behaviors than non-Facebook users. It is unclear whether these biases that are unaccounted for would have a systematic or random effect on our results. However, we do expect their impact to be relatively small, particularly as our ‘ground-truth’-based debiasing approach adjusts masking estimates regardless of the source of non-response bias, and as supported by our community-reported masking analysis (since the community question captures some of the non-Facebook user population). We assumed that self-reported mask-wearing is biased in the same magnitude and direction as self-reported COVID-19 vaccination status – an assumption that should be tested in future research. Our approach also does not resolve issues of small sample size; for example, the association we found between bias-corrected self-reported masking and community-reported masking is stronger between modeled estimates than between raw means. Although we have attempted to correct for bias in our mask-wearing estimates, the point estimates for these values should be interpreted with caution. The goal of our work isn’t to produce point estimates of county-level mask-wearing behavior but instead to take advantage of the CTIS survey design and characterize the relative trends in mask-wearing behavior between and across counties. We advocate for additional observational studies with experimental designs that would allow for direct estimation of these quantities to improve behavioral surveillance estimates.

In summary, we have produced the first accurate high-resolution spatiotemporal estimates of face mask wearing in the United States for the period from September 2020 through May 2021. Our work reveals that masking behavior is highly variable across the United States, suggesting that a one-size-fits-all approach to increasing mask wearing behavior is likely to be ineffective. Instead, we have identified regions of the country with higher and lower masking levels. These differences should be investigated going forward as public health organizations consider how to more effectively target these low masking regions. For example, these communities may be more susceptible to mis- and disinformation regarding mitigation behaviors which must be strategically confronted. Furthermore, this variability in behavior demonstrates the need to develop infectious disease dynamics models to analyze and predict how spatiotemporal trends in disease are affected by changes in human behavior, such as vaccination, contact patterns, and face mask-wearing. Our analyses also address issues of survey bias, with the takeaway that, in the future, we should invest in robust survey infrastructure that can recruit large representative samples with minimal bias, including using certain representative respondents as human social sensors to report on their communities.

## Data Availability

Data aggregated to the county-month scale and all code to analyze these data will be made available on GitHub at https://github.com/bansallab/spatial_masking.

## Acknowledgments

The authors thank the Carnegie Mellon University Delphi team for sharing the US COVID-19 Trends and Impact Survey openly and freely, and Alex Reinhart for his feedback on this work. We also thank the OutbreaksNearMe team at Boston Children’s Hospital and Momentive for data sharing, and Benjamin Rader and John Brownstein for their feedback.

Research reported in this publication was supported by the National Institute of General Medical Sciences of the National Institutes of Health under award number R01GM123007. The content is solely the responsibility of the authors and does not necessarily represent the official views of the National Institutes of Health.

## Authors’ contributions

JCT performed analyses, interpreted results, drafted, and edited the manuscript. ZS designed and guided analyses, and edited the manuscript. SB designed the study, guided analysis, interpreted results, and edited the manuscript. All authors read and approved the final manuscript.

## Data availability

Data aggregated to the county-month scale and all code to analyze these data will be made available on GitHub at https://github.com/bansallab/spatial_masking. Individual survey responses cannot be shared by the authors, but researchers can visit https://cmu-delphi.github.io/delphi-epidata/symptom-survey/data-access.html if they would like to enter an agreement for data usage with CMU Delphi.

## Supplementary Materials

### Additional CTIS details

The COVID-19 Trends and Impact Survey was created by researchers in the Delphi Group at Carnegie Mellon University and distributed via Facebook to active users 18 or older starting in April 2020. On a daily basis, a random state-stratified sample of Facebook users are invited to take the survey at the top of their news feed. These users will not be re-invited to take the survey for at least thirty days. The survey asks a broad range of questions related to COVID-19 symptoms and behaviors, with variations across each of the thirteen different waves spanning from April 2020 to June 2022. In this study, we make use of data from Wave 4 (September 8, 2020 - November 23, 2020), Wave 5 (November 24, 2020 - December 18, 2020), Wave 6 (December 19, 2020 - January 11, 2021), Wave 7 (January 12, 2021 - February 7, 2021), Wave 8 (February 8, 2021 - March 1, 2021), and Wave 10 (March 2, 2021 - May 19, 2021, Wave 9 was skipped for numbering purposes).

Weights were provided by Facebook for each response to adjust for non-response and coverage bias at the daily-state level [58]. Briefly, each weight describes the number of people represented by a respondent based on their age, gender, location, and date of response. These weights were calculated via a two-step process using inverse propensity score weighting based on respondents’ demographics recorded in their Facebook user profiles to adjust the survey sample to reflect active Facebook users, followed by revisions of these weights using post-stratification so that the survey sample reflects the general population [27]. Because these weights were not at the same scale of our data analysis, we did not use them and instead performed raking to calculate our own weights as described in Methods.

As part of our data processing, we also dropped responses missing fips codes or outside the 50 states and the District of Columbia. When raking, we use three categories for age (18-24, 25-54, and 55+), two categories for sex (ACS)/gender (CTIS) (male and female), and six categories for education (less than high school, high school graduate or equivalent, some college, 2 year college degree, 4 year college degree, and postgraduate degree). Subsetting age further results in issues of nonconvergence. Raking on race/ethnicity with more than two categories leads to substantial nonconvergence whether or not we include education. We drop responses missing age, gender, or education from being raked.

In addition, when dichotomizing CTIS responses we assume that people who “sometimes” wore a face mask in public were not masking which may lead us to underestimate self-reported masking levels. We believe the effects of this assumption are minimal because the proportion of “sometimes” responses was small compared to other options (Figs. S14).

### Estimation of CTIS bias

We compare CTIS responses about vaccination to true vaccination estimates for the period from April 1, 2021 through May 31, 2021. We chose this period when nearly all adults were eligible for vaccination in the U.S. so that the sample population responding to the masking and vaccination questions would be most similar. In this time period, the differences between the survey and ground-truth vaccination data have also stabilized (Fig. S18). We use a (frequentist) binomial generalized linear mixed-effects model to estimate *p*_*i*_, the proportion of respondents who were vaccinated at the county-level each week. If *V*_*i*_ is the number of (partially) vaccinated respondents in each county *i* out of *N*_*i*_ respondents, then the model is as follows:

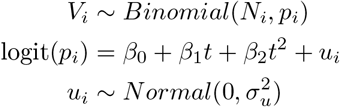

where *t* and *t*^2^ are orthogonal polynomials of degree 1 and 2, respectively, generated by the `poly` function in R from the rank of the weeks in which the vaccination data were observed. Therefore, *β*_1_ and *β*_2_ are covariates that describe the trend of time in expected reported vaccination across counties, while *u*_*i*_ describes systematic difference in vaccination in county *i* relative to the mean trend. This model was implemented using `glmer` in the `lme4` package [36].

Using these modeled CTIS county-level vaccination proportions, we compared them with the true vaccination data to calculate the expected bias in reported survey responses relative to ground truth data in county *i*. There were 45 counties that had missing bias estimates due to either a lack of weekly CTIS vaccination survey responses between 0 and 1 (cannot use logits of *p* = 0, 1), or missing true vaccination estimates for weeks with survey responses between 0 and 1 (therefore, cannot calculate difference between true and CTIS vaccination estimates).

### CTIS model coefficients

Model coefficients for z-score(log_10_(population density)) ranged from 0.45 to 0.55 (Fig. S22), meaning a one unit change in z-score(log_10_(population density)) is correlated with the expected odds of masking multiplying by *e*^0.5^ ≈ 1.65. The coefficient of the population density covariate in our binomial regression model is consistent over time, indicating that the relationship between population density and masking behavior is stable across months.

### CTIS mixed effects model specifications

In addition to the models presented in the main text, we ran two models to estimate mask-wearing at the county-month level using state or county-level random effects. For both models, we define *M*_*i*_ as the number of respondents masking in county *i* (e.g., respondents that masked most or all of the time in the past 5-7 days), *N*_*i*_ as the total number of respondents in county *i* (*M*_*i*_ ≤ *N*_*i*_), and *p*_*i*_ as the county-level probability of a response consistent with masking. We ran both models using `brms` [29] with the `cmdstanR` [30] backend and we ran the sampler with 4 chains for 3000 iterations per chain. We use the following model to estimate 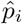 and 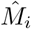 with state-level effects

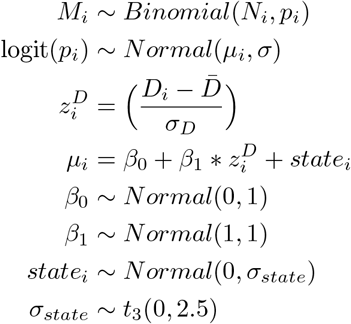

where *D*_*i*_ = log_10_(`population density`_*i*_) for county *i*. The 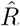 values were frequently ≥ 1.02 and *n*_*eff*_ *<* 500 for the intercept population-level effect and group-level effects, indicating lack of convergence. The population density coefficient did show convergence (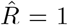 and *n*_*eff*_ *>* 1800) and remained consistent over time and close to the values observed in the original model. Only approximately 1% of observations had Pareto k values *>* 0.7 indicating that the model was robust to the influence of individual observations.

For county-level effects we used the following model to estimate 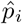 and 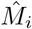:

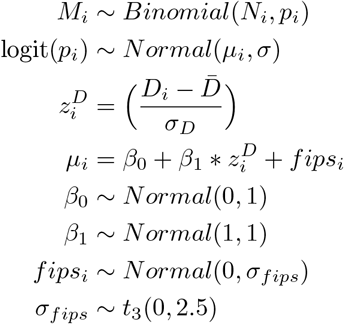

where *D*_*i*_ = log_10_(`population density`_*i*_) for county *i*. In this case sampler diagnostics indicated reasonable model convergence with 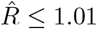 and *n*_*eff*_ *>* 600 for all months except April and May 2021 when *n*_*eff*_ *>* 250. However, over 50% of observations in each month had Pareto k values *>* 0.7, indicating the influence of each data point and potential overfitting. As observed with the state-level random effects model, the population density coefficient remained consistent over time with values near those from the original model.

### Outbreaks Near Me survey details & results

The Outbreaks Near Me (ONM) survey was created by scientists at Harvard and Boston Children’s Hospital and distributed through a partnership with SurveyMonkey. Following the completion of another survey on SurveyMonkey, a random representative sample of users across the United States were invited to take the Outbreaks Near Me survey. The survey was released in June 2020 and asked respondents how likely they were to wear a mask in several different environments: while grocery shopping, visiting with friends and family in their homes, exercising outside, and in the workplace. Answer options were (1) Very, (2) Somewhat, (3) Not so, or (4) Not likely at all (Fig. S23). We focus on the responses to the grocery shopping scenario, as this setting is most comparable to the “in public” scenario described in the CTIS question. To dichotomize these responses for an analysis of the proportion of respondents wearing masks, we consider “very likely” responses as masking and all other responses as not masking. This choice allows for comparison with CTIS data and makes sense given the small percentage of “somewhat likely” responses (Fig. S24). We aggregate responses at the zipcode-month scale and crosswalk these estimates to the county-month level using HUD files [59]. The Outbreaks Near Me survey dropped responses from individuals who reported their age as less than 13 or more than 100 years old. We additionally drop respondents who did not respond to the grocery store portion of the masking question or who have an invalid zip code that cannot be crosswalked to a fips code; this process leaves us with 1,042,685 valid responses. Survey weights were provided for individual responses at the weekly-state and daily-national scales, though we do not to use them because our analysis focuses on the county-month scale.

Due to small sample sizes, we use binomial regression models to estimate masking proportions for each county-month, as described in ‘Bayesian binomial regression model’ in the Methods (Figs. S25, S26). We compare estimates from these models to CTIS values calculated using only the binomial regression model, i.e., no raking/resampling or debiasing. We calculate the ratio of ONM to CTIS masking proportions for each county each month and take the average across all months in the survey, excluding counties that have estimates for fewer than five of nine months. Additionally, we visualize the time series of individual counties from the two surveys side by side.

**Figure S1.**
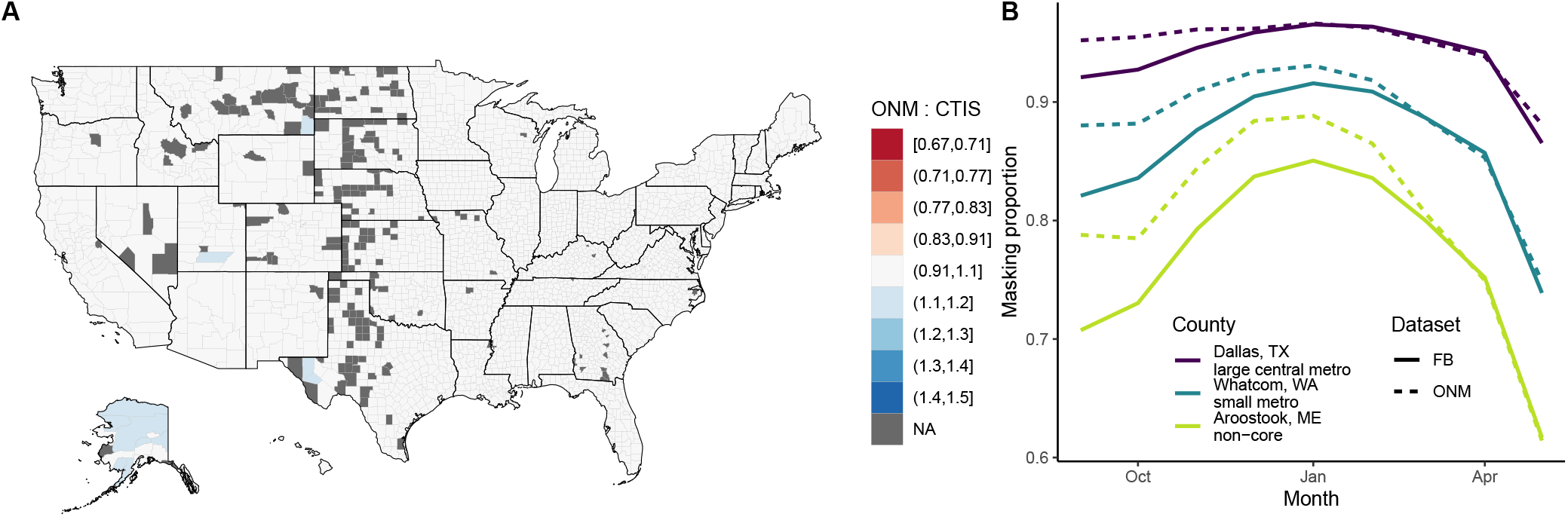
CTIS and ONM responses show similar values and trends across space and time. (A) Average ratio of ONM to CTIS masking proportions for each county from September 2020 through May 2021. All average ratios are greater than 1, but only 0.4% are greater than 1.1. Counties with estimates for less than five of nine months are excluded. (B) Across three counties of varying urbanicity, masking proportions increase through January 2021 and then decrease through May 2021 in both surveys. Differences between survey estimates appear to decrease over time. Urban counties may exhibit higher levels of masking and less variability in these estimates over time compared to rural counties. Both CTIS and ONM estimates are outputs from binomial regression models without raking or bias offsets.

**Figure S2.**
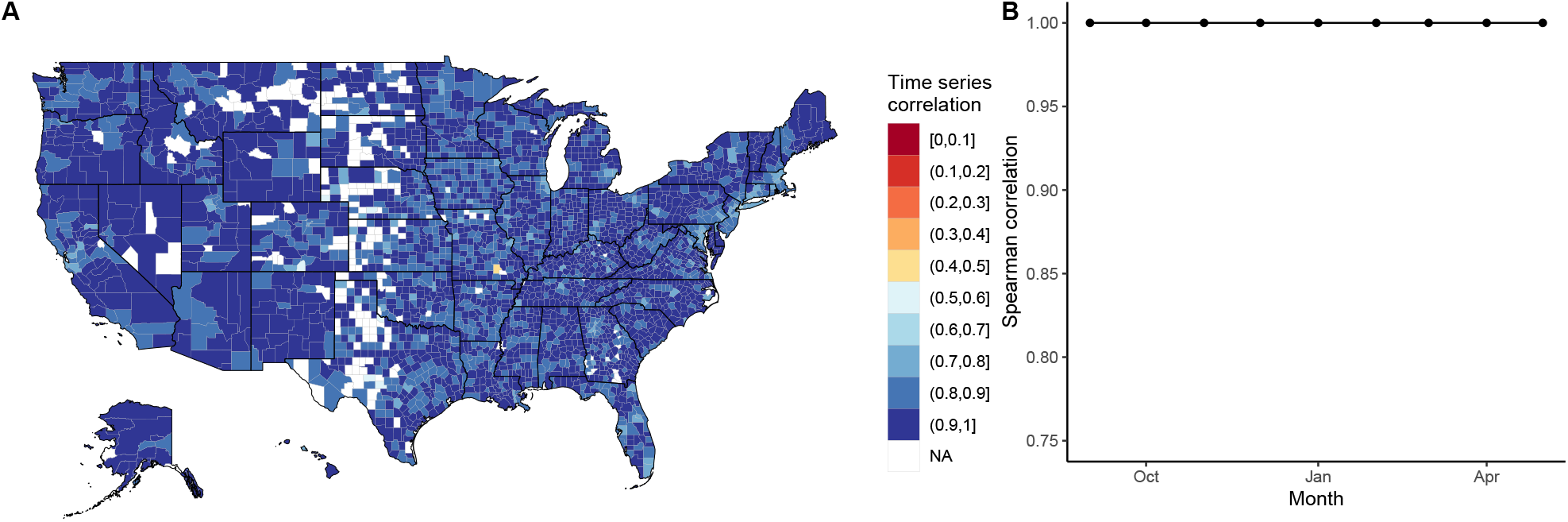
Time series correlations between the CTIS and ONM surveys. (A) Time series correlation for each county across all months is high across all counties. (B) Time series correlation for all counties each month is 1 across all months. Both CTIS and ONM estimates are outputs from binomial regression models without raking or bias offsets.

**Figure S3.**
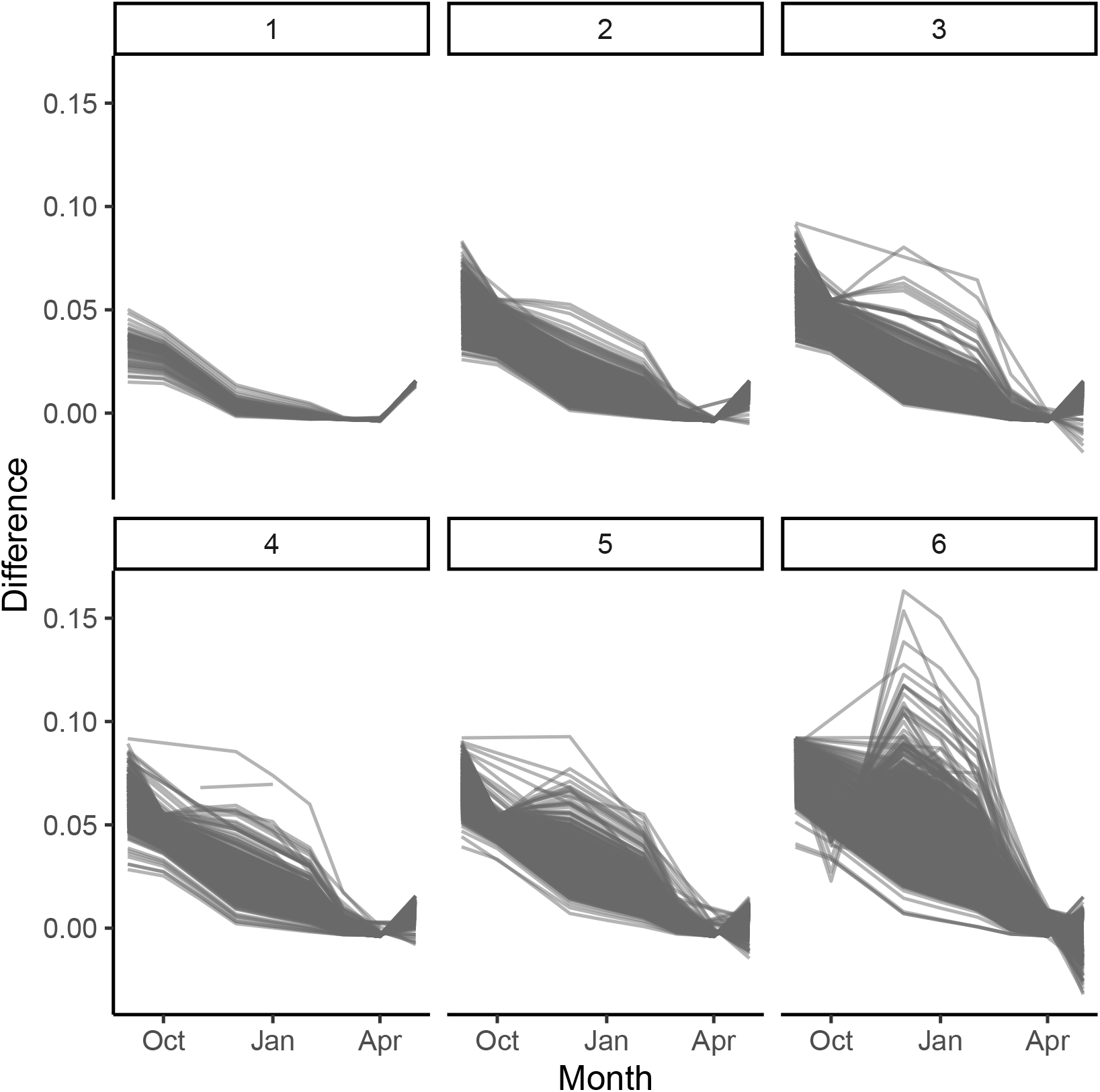
Difference between county masking estimates from ONM and CTIS surveys decreases over time, with greater variability in more rural counties. Facets designate NCHS urban-rural classification with 1 being the most urban and 6 the most rural. Difference between survey estimates goes to nearly 0 across all counties in April 2021, with increasing, albeit small, differences in May 2021. Both CTIS and ONM estimates are outputs from binomial regression models without raking or bias offsets.

**Figure S4.**
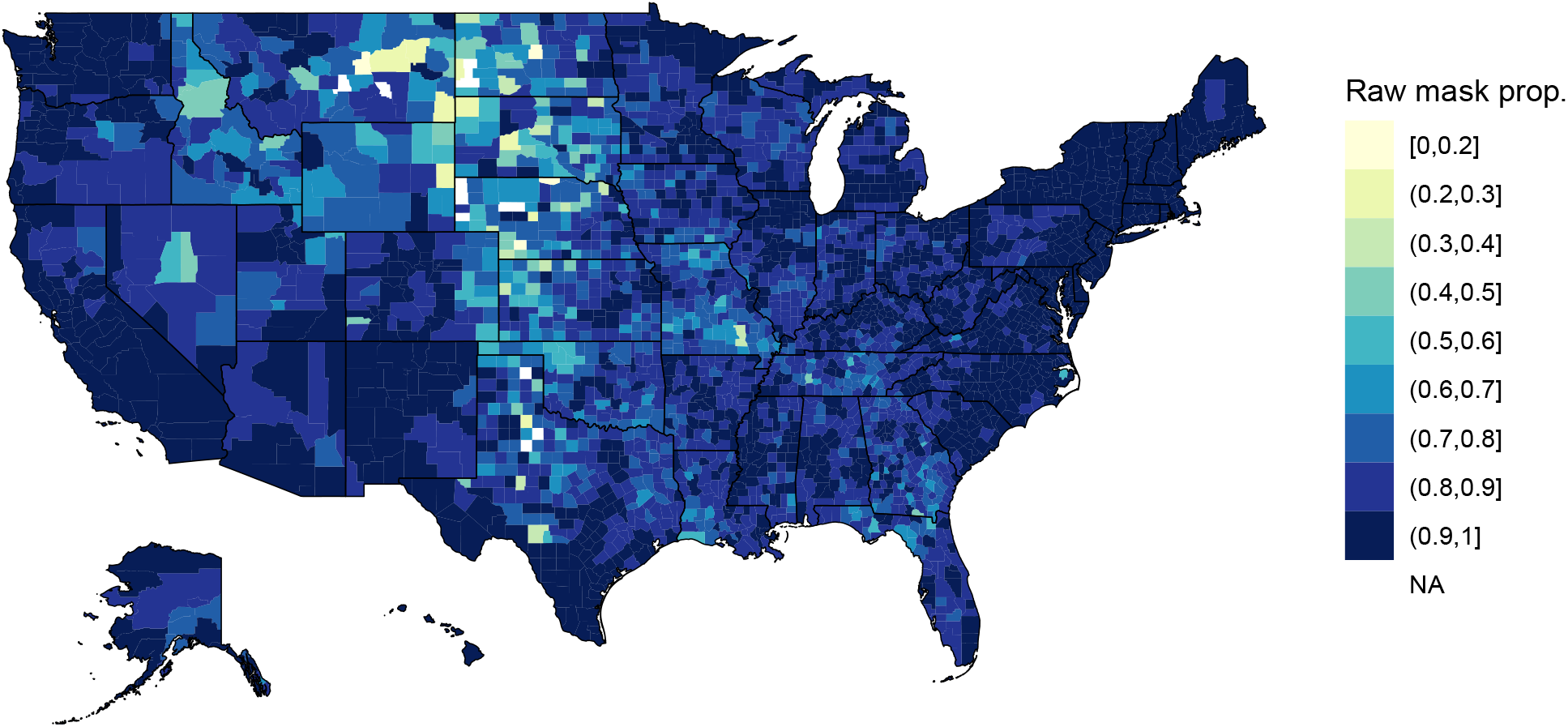
Raw CTIS masking data for February 2021. Proportions masking most of the time or more are much higher than observational studies would suggest, with spatial trends difficult to identify.

**Figure S5.**
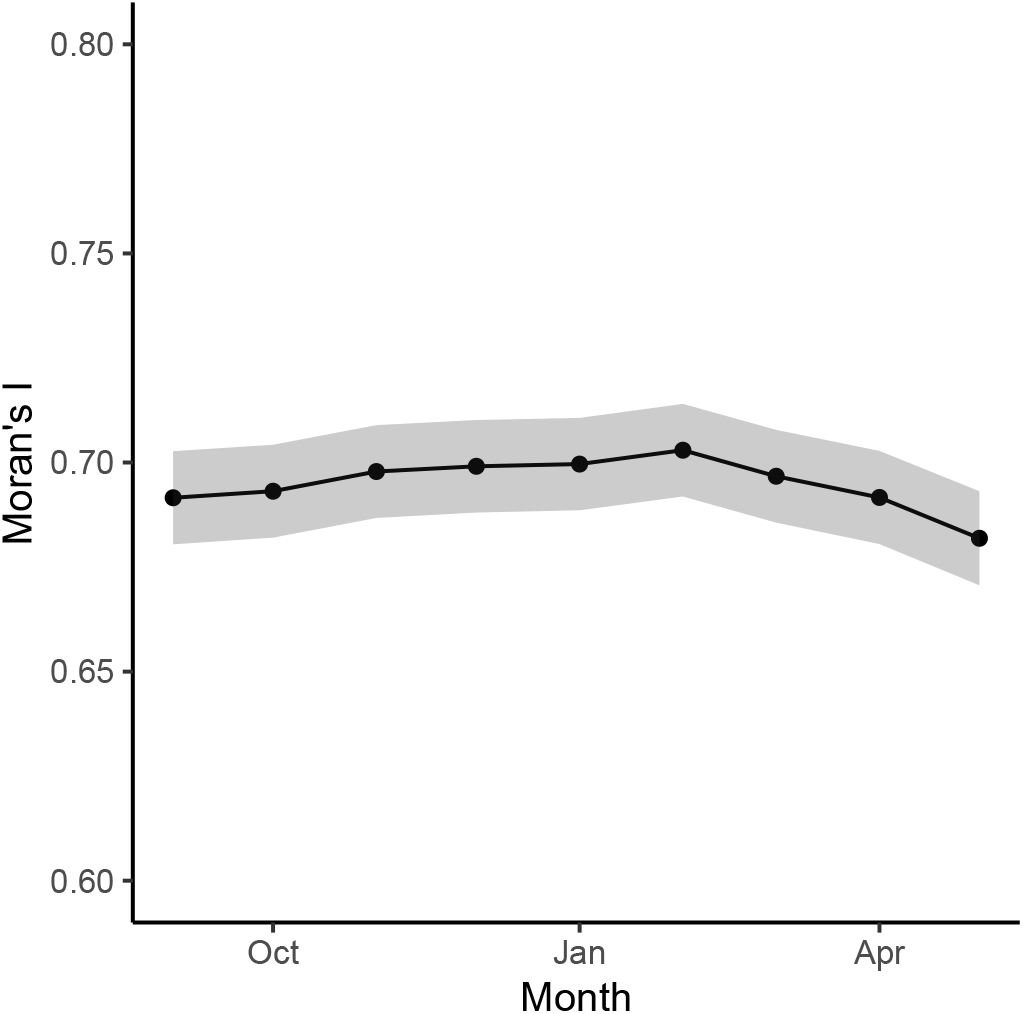
Spatial heterogeneity in self-reported masking behavior does not vary over time. Moran’s I calculated for self-reported masking behavior from the CTIS estimates from binomial regression model with raking/resampling and bias offset.

**Figure S6.**
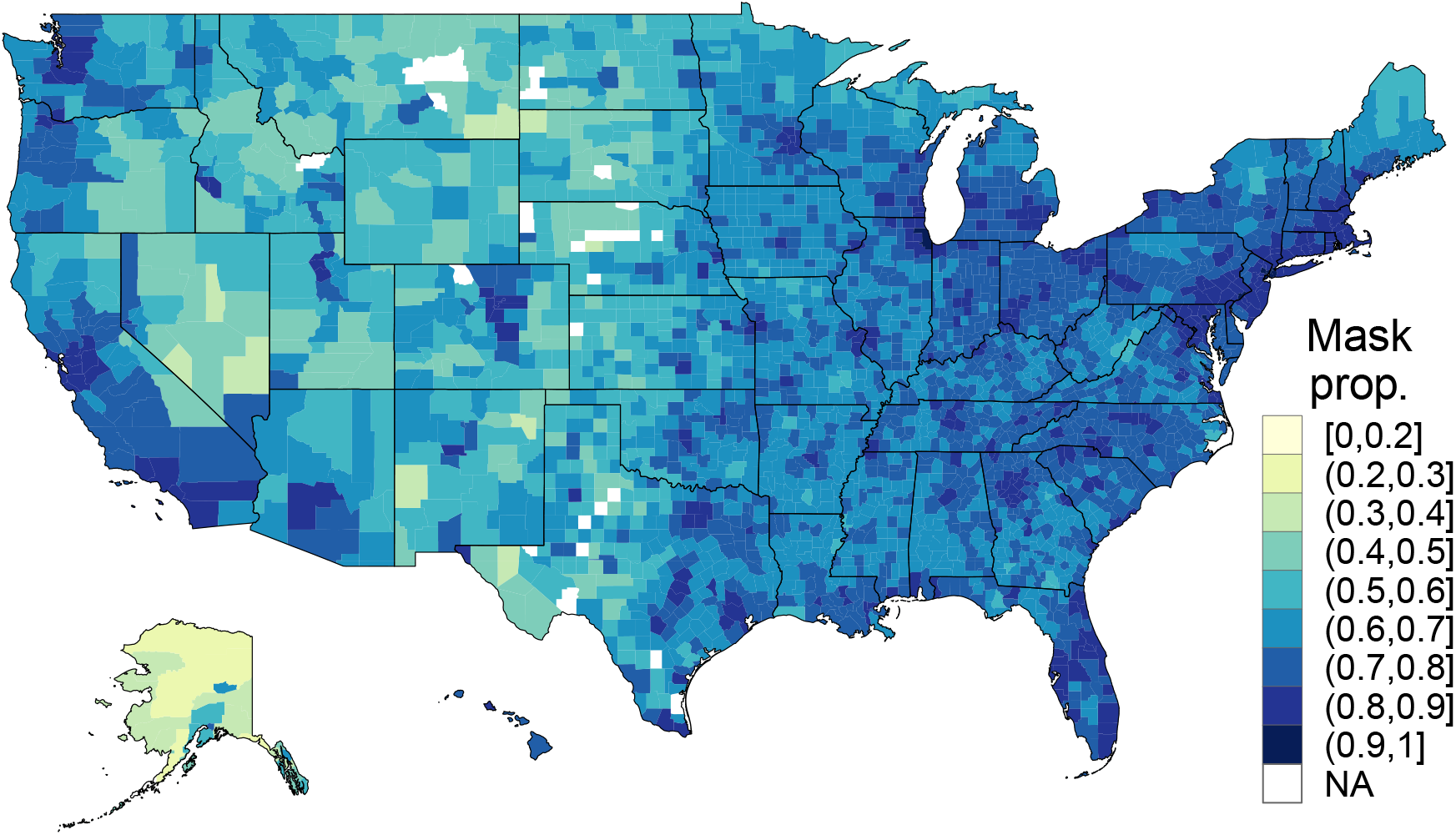
Bias-corrected masking estimates for December 2020.

**Figure S7.**
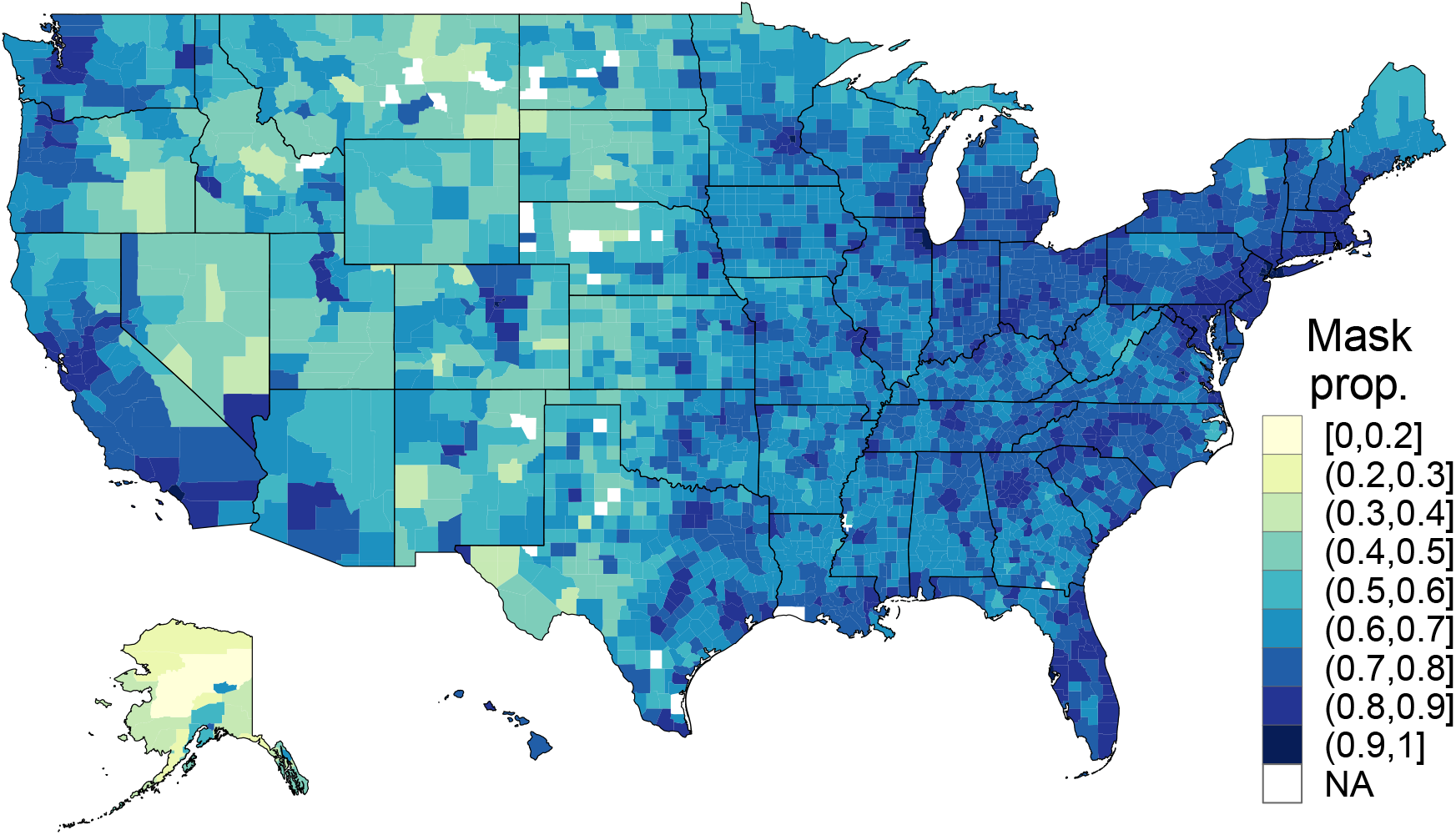
Bias-corrected masking estimates for February 2021.

**Figure S8.**
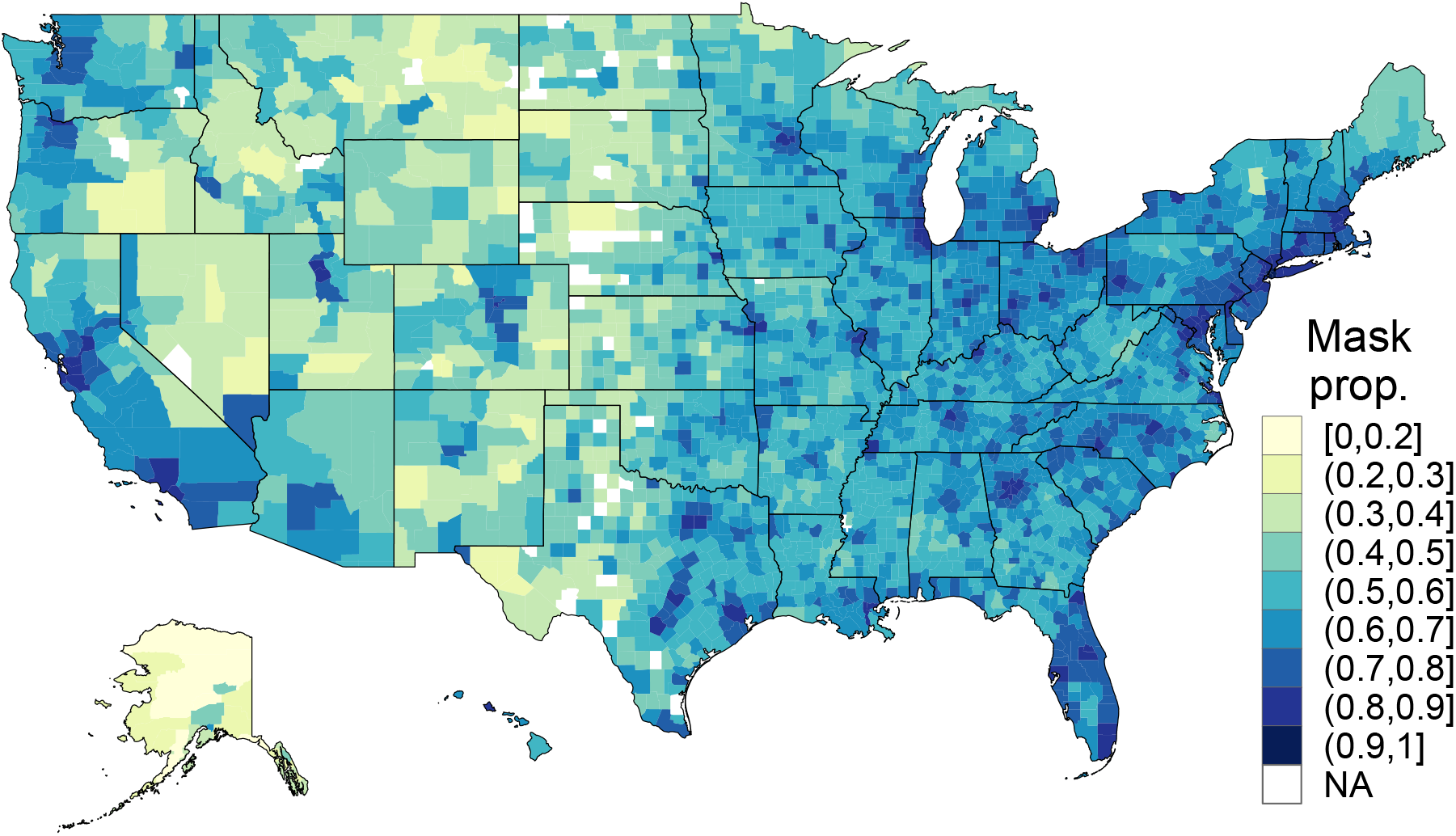
Bias-corrected masking estimates for April 2021.

**Figure S9.**
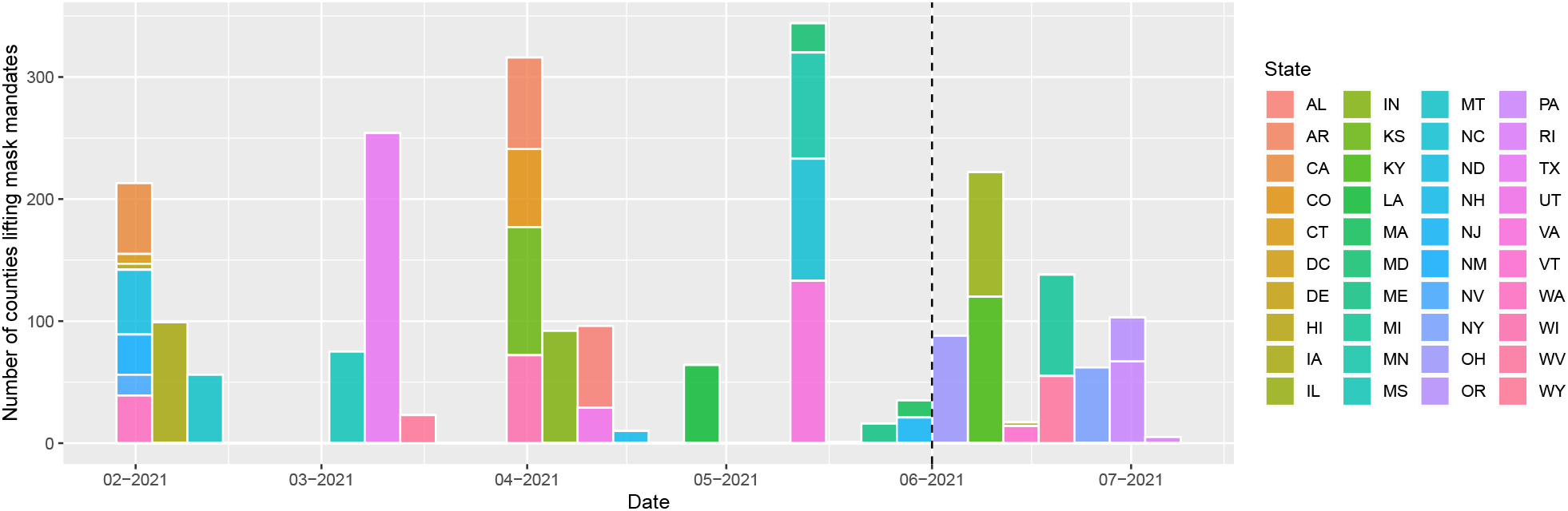
Timeline of lifting of county mask mandates in the U.S. Neither state nor county mask mandates were ever imposed in AK, AZ, FL, GA, ID, MO, NE, OK, SC, SD, and TN during the period from April 10, 2020 to August 15, 2021. Thus, these states have been excluded. Approximately 49% of counties that ever imposed a mask mandate lifted it before May 1, 2021.

**Figure S10.**
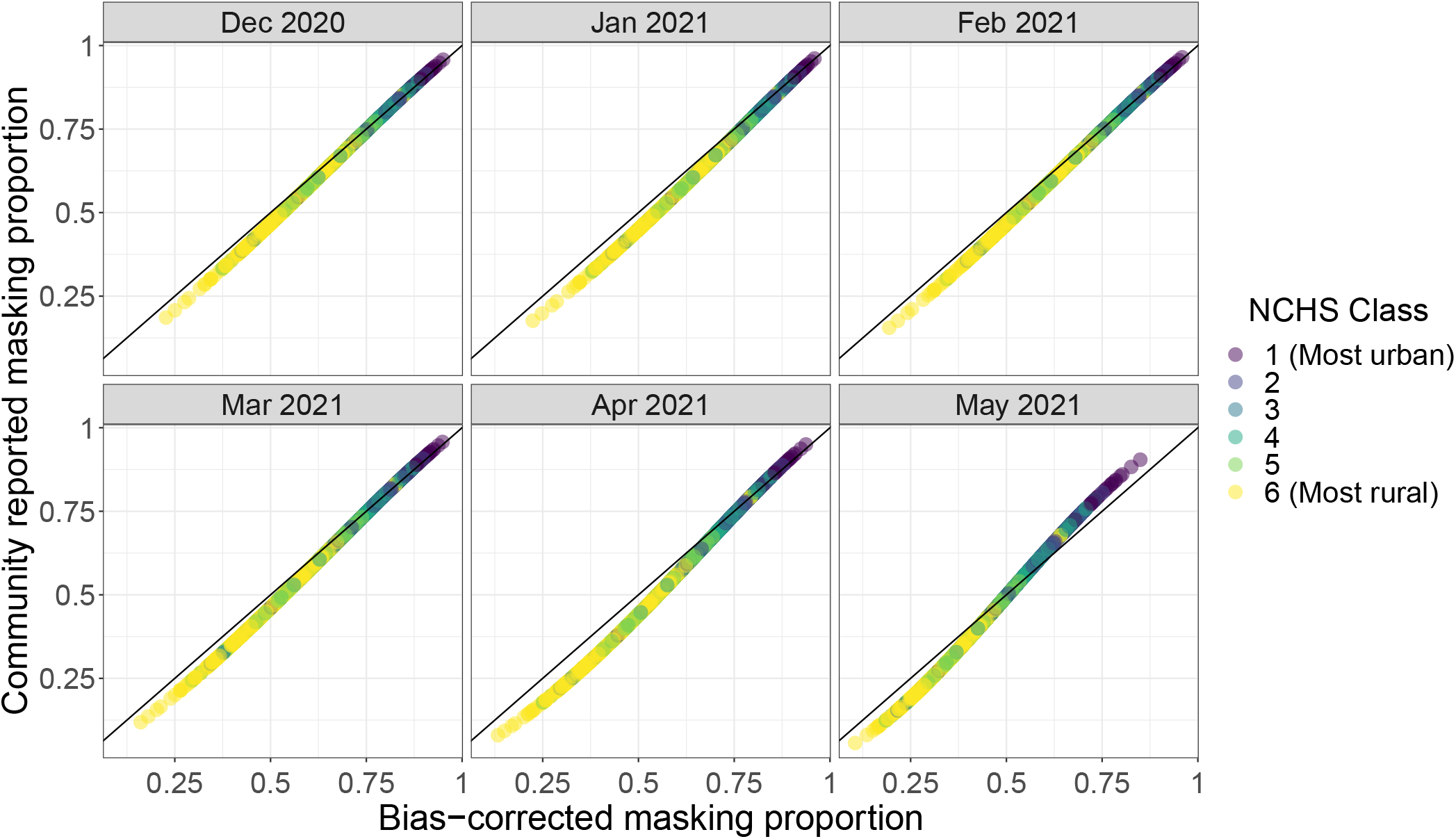
Community reported masking gives a good estimate of bias-corrected self-reported masking even when influential fips code are removed from the model. Fips codes with pareto k values *>*= 0.7 were excluded from the Bayesian binomial regression model with bias offsets (specifically fips 4019, 6037, 6071, 12071, 12103, 36005, 40143, 41039, 45045, 48201, 48439, 53033). Community reported masking refers to the CTIS question where individuals report how many people in their community are masking, which may decrease non-response and social desirability bias compared to asking individuals to self-report their masking behavior. Point color denotes urban-rural classes.

**Figure S11.**
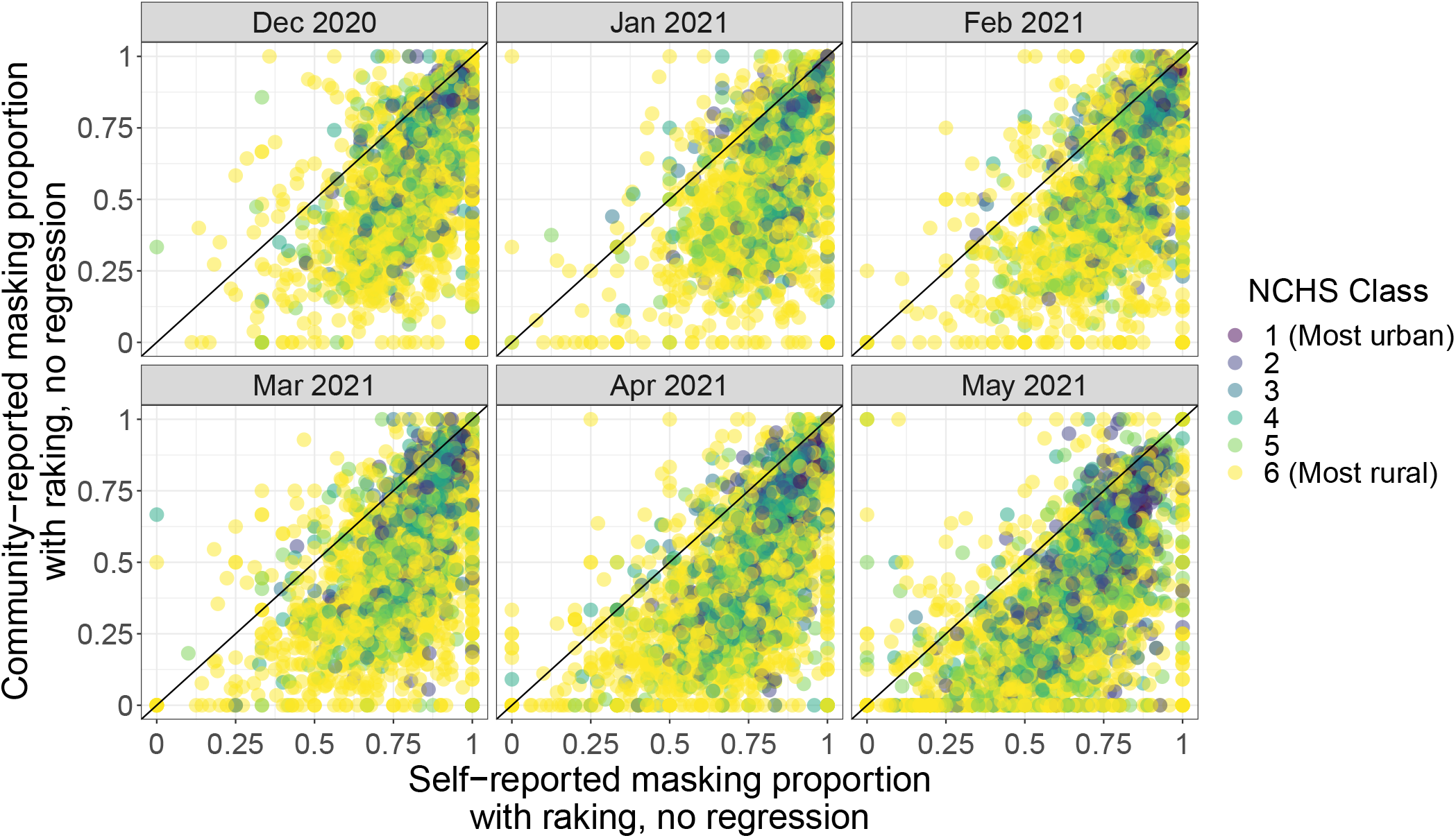
Self-reported masking estimates generally exceed community-reported estimates without bias-correction, but the data are noisy without the binomial regression model. Both masking estimates are calculated from raked and resampled observations but are not run through a binomial regression model to correct for small sample size. Additionally, self-reported masking estimates are not debiased. Recall that community reported masking refers to the CTIS question where individuals report how many people in their community are masking, which may decrease non-response and social desirability bias compared to asking individuals to self-report their masking behavior. Point color denotes urban-rural classes.

**Figure S12.**
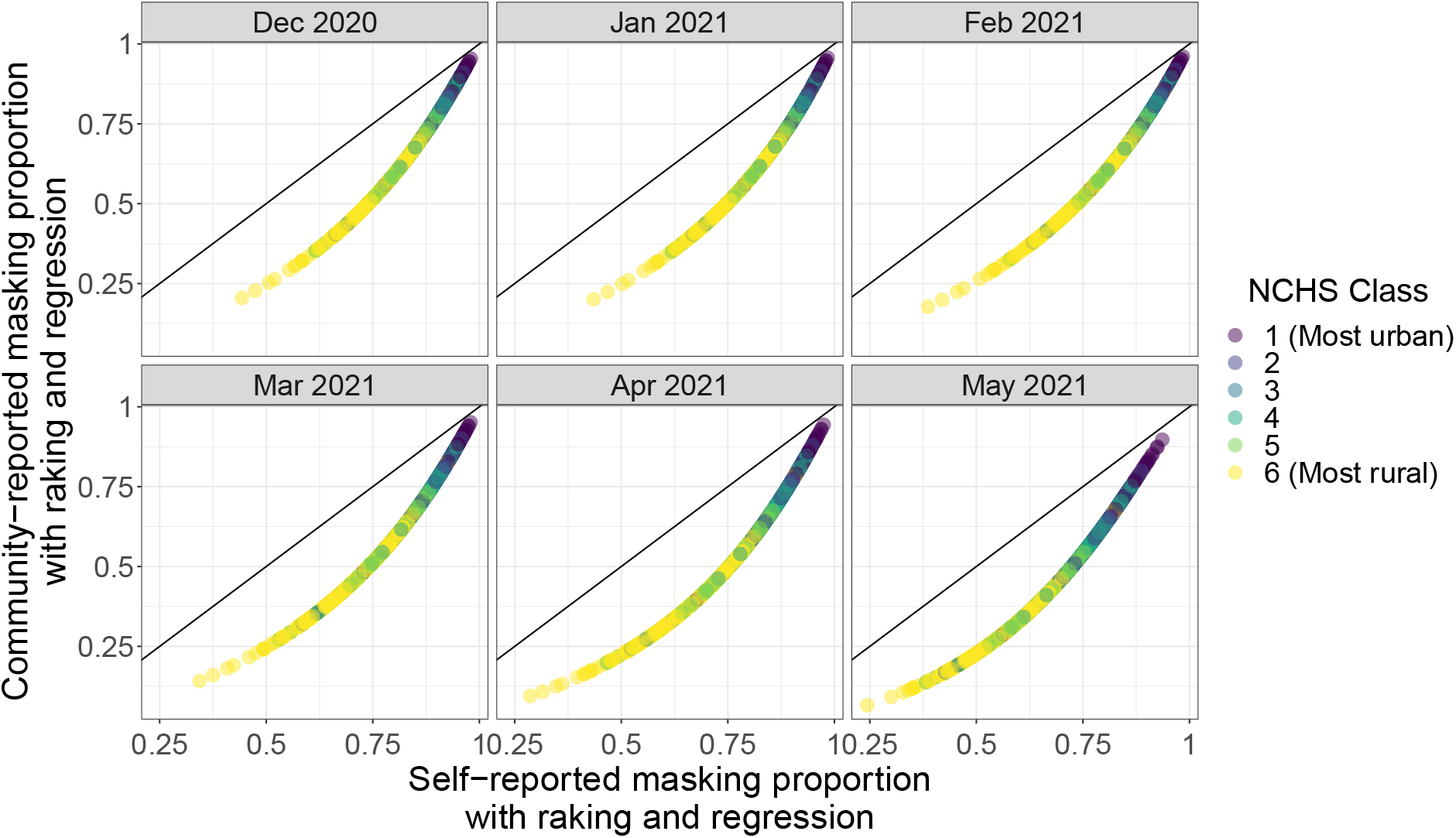
Non-bias-corrected self-reported masking estimates exceed community-reported estimates to a greater degree in rural counties. Both masking estimates are calculated from raked and resampled observations run through a binomial regression model to correct for small sample size. Self-reported masking estimates are not debiased. Recall that community reported masking refers to the CTIS question where individuals report how many people in their community are masking, which may decrease non-response and social desirability bias compared to asking individuals to self-report their masking behavior. Point color denotes urban-rural classes.

**Figure S13.**
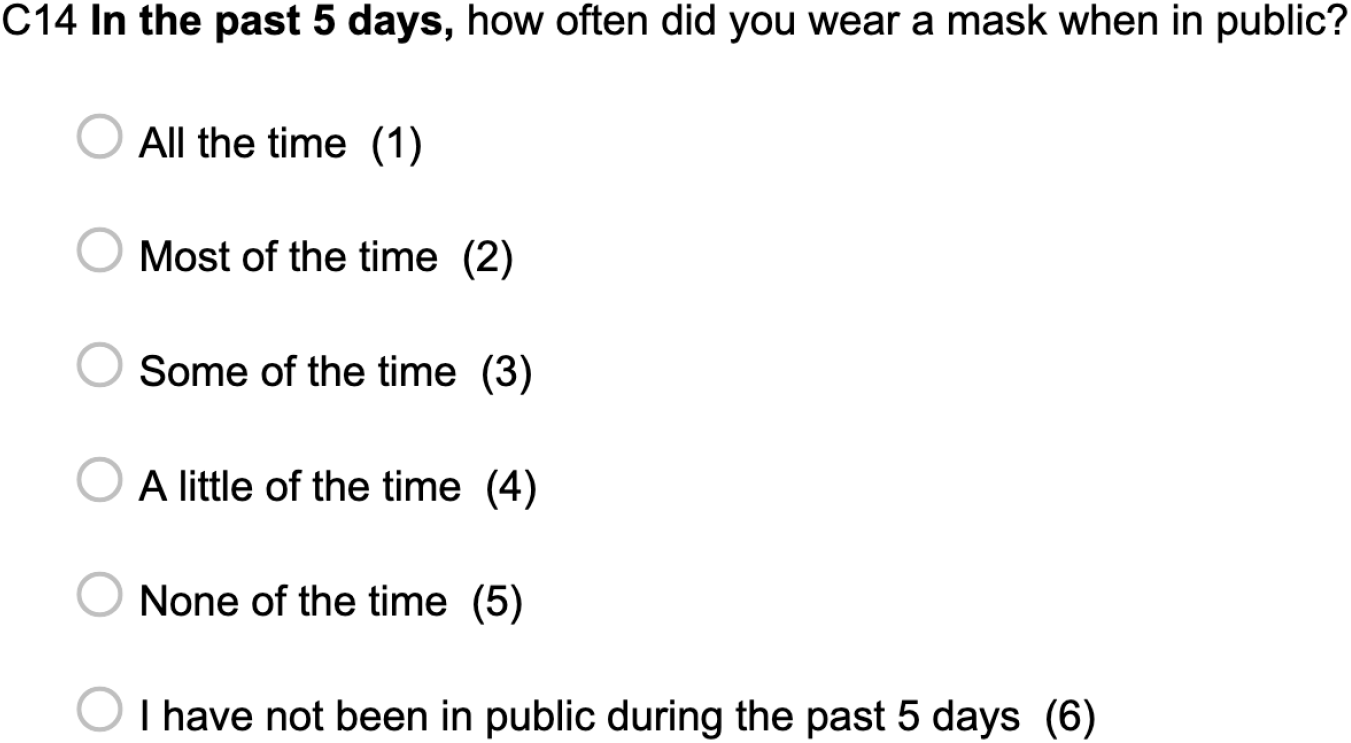
CTIS question on masking behavior. Note that time period of interest changed from 5 days to 7 days on February 8, 2021.

**Figure S14.**
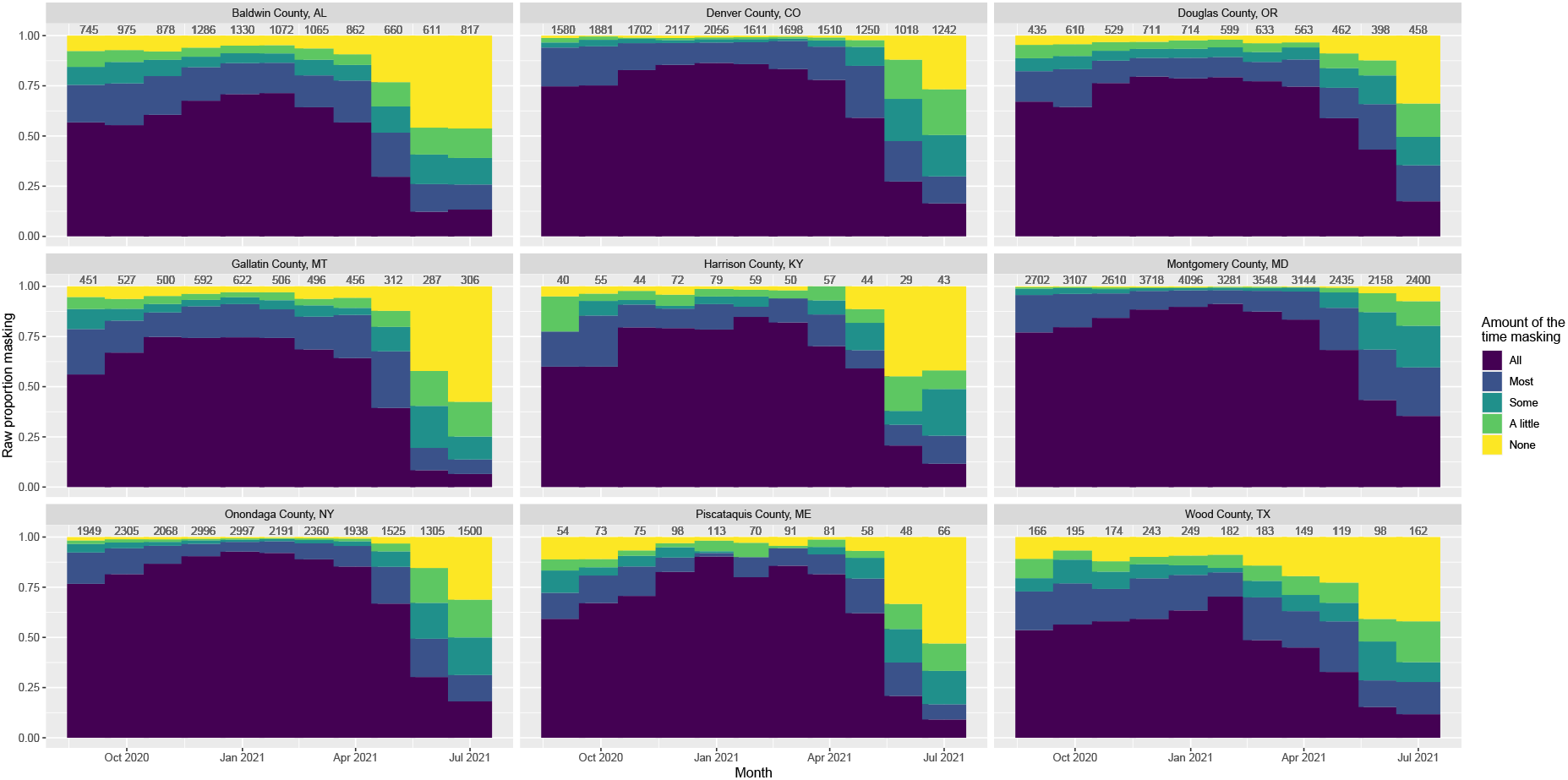
Raw proportions of each CTIS response for a selection of counties spanning population densities across time. Number of responses in that county-month listed above each bar. NCHS urban-rural classifications are as follows: (1) Denver, CO; (2) Montgomery, MD; (3) Onondaga, NY; (4) Baldwin, AL; (5) Gallatin, MT; Douglas, OR; (6) Wood, TX; Piscataquis, ME; Harrison, KY.

**Figure S15.**
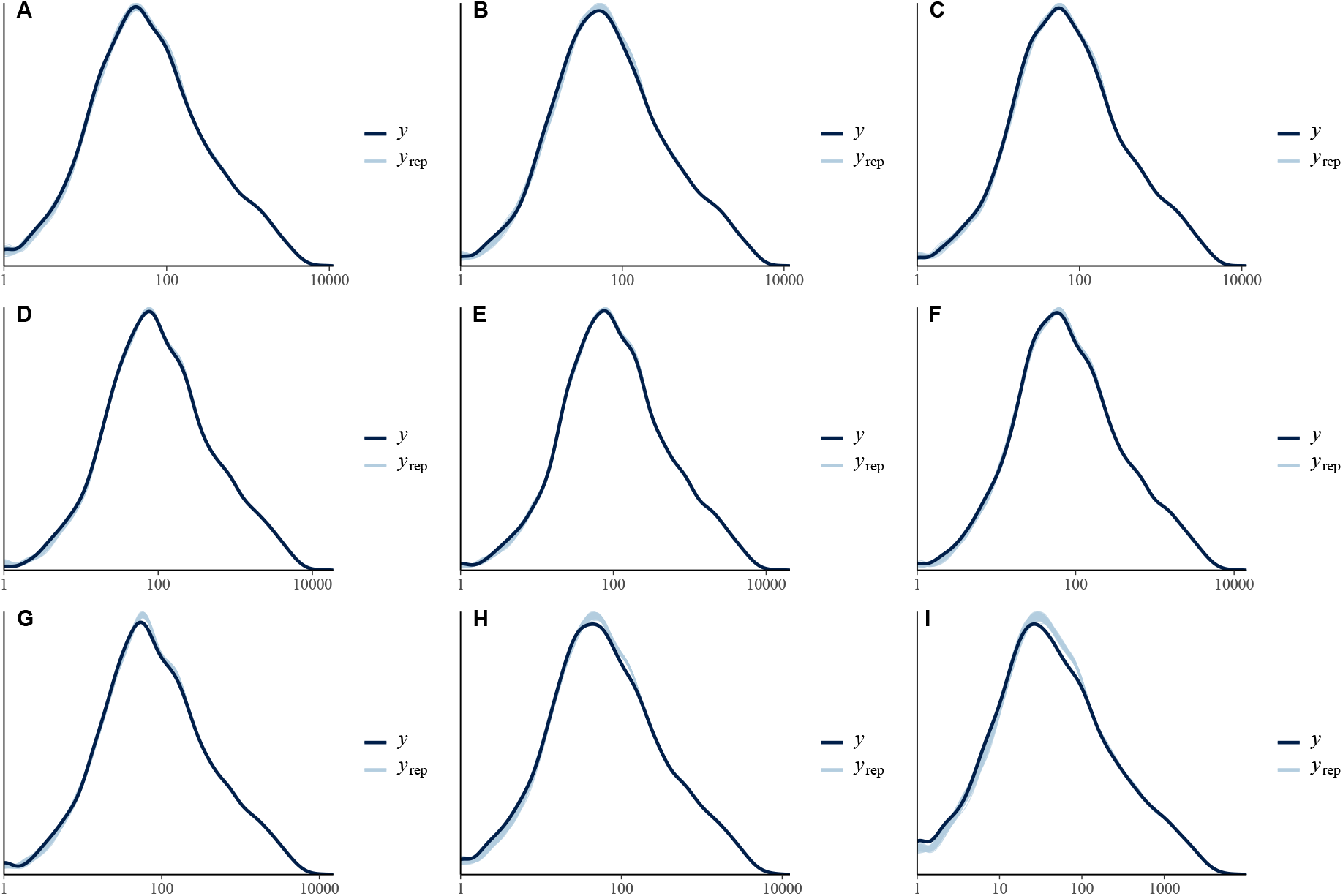
Posterior predictive checks of binomial regression model of CTIS data (no raking or bias offset) with x-axis on a log-10 scale for visual aid. These figures compare observed data (*y*, black) to model predictions (*y*_*rep*_, blue). The predictions well-approximate the observed data with no systematic differences indicating that the model fit is reasonable.

**Figure S16.**
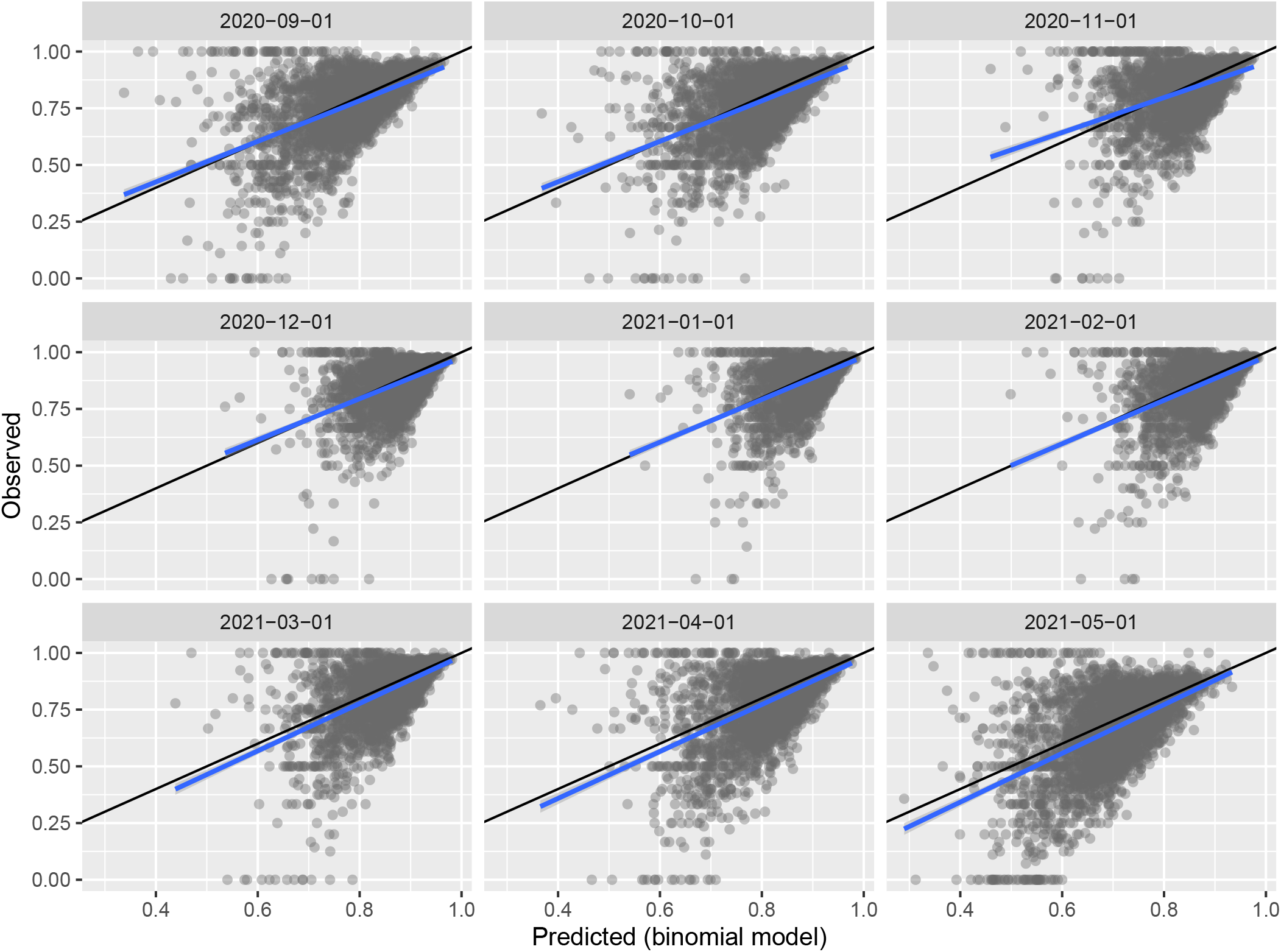
Observed versus predicted CTIS masking estimates with binomial regression model (no raking or bias offset). Black line shows y = x; blue line is a linear fit of the data. The linear trend is close to the one-to-one line indicating good model fit, though there is substantial noise.

**Figure S17.**
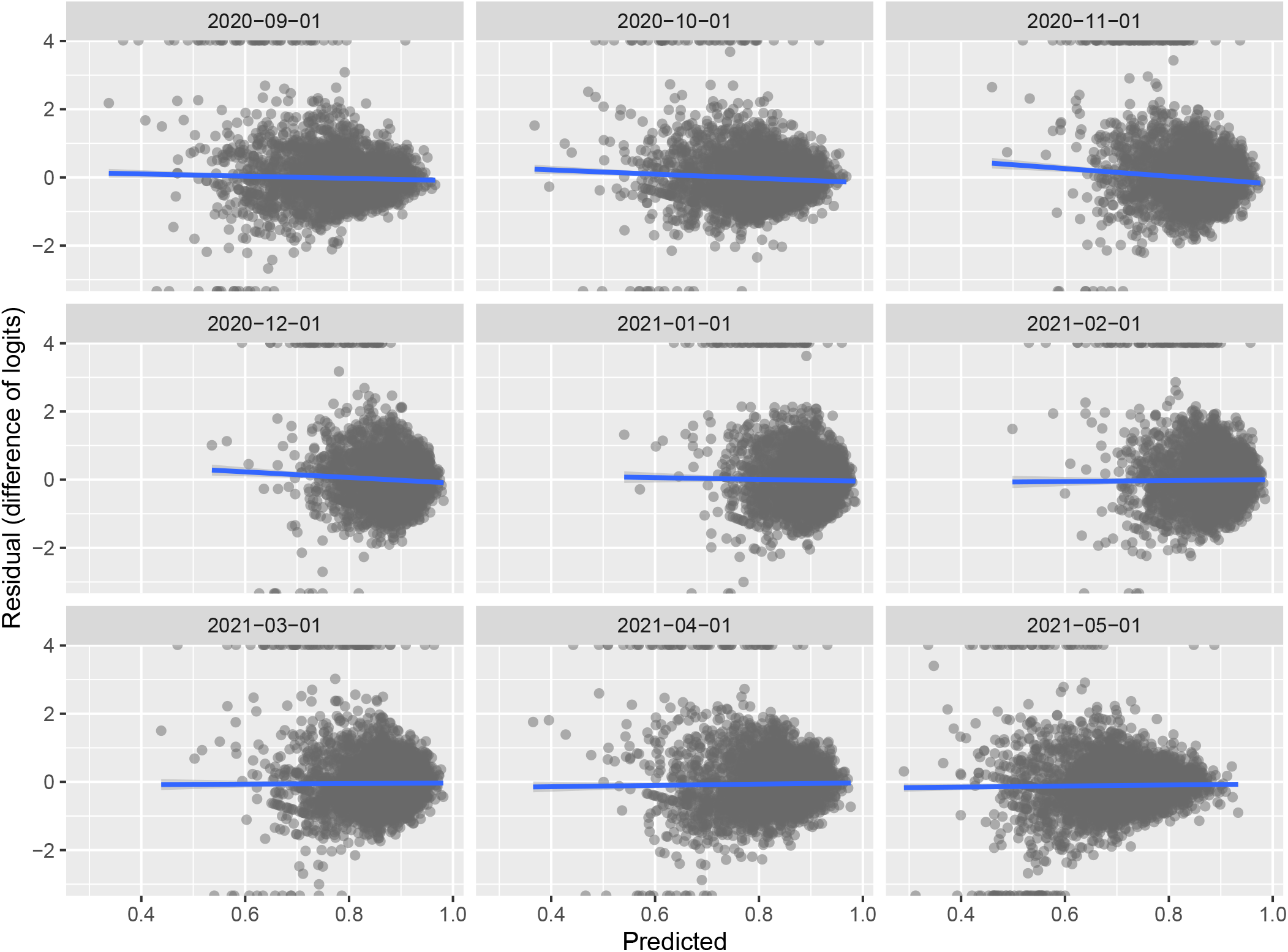
Observed versus residual CTIS masking estimates with binomial regression model (no raking or bias offset). Blue line is a linear fit of the data. Residuals show little association with predicted value which indicates a reasonable model.

**Figure S18.**
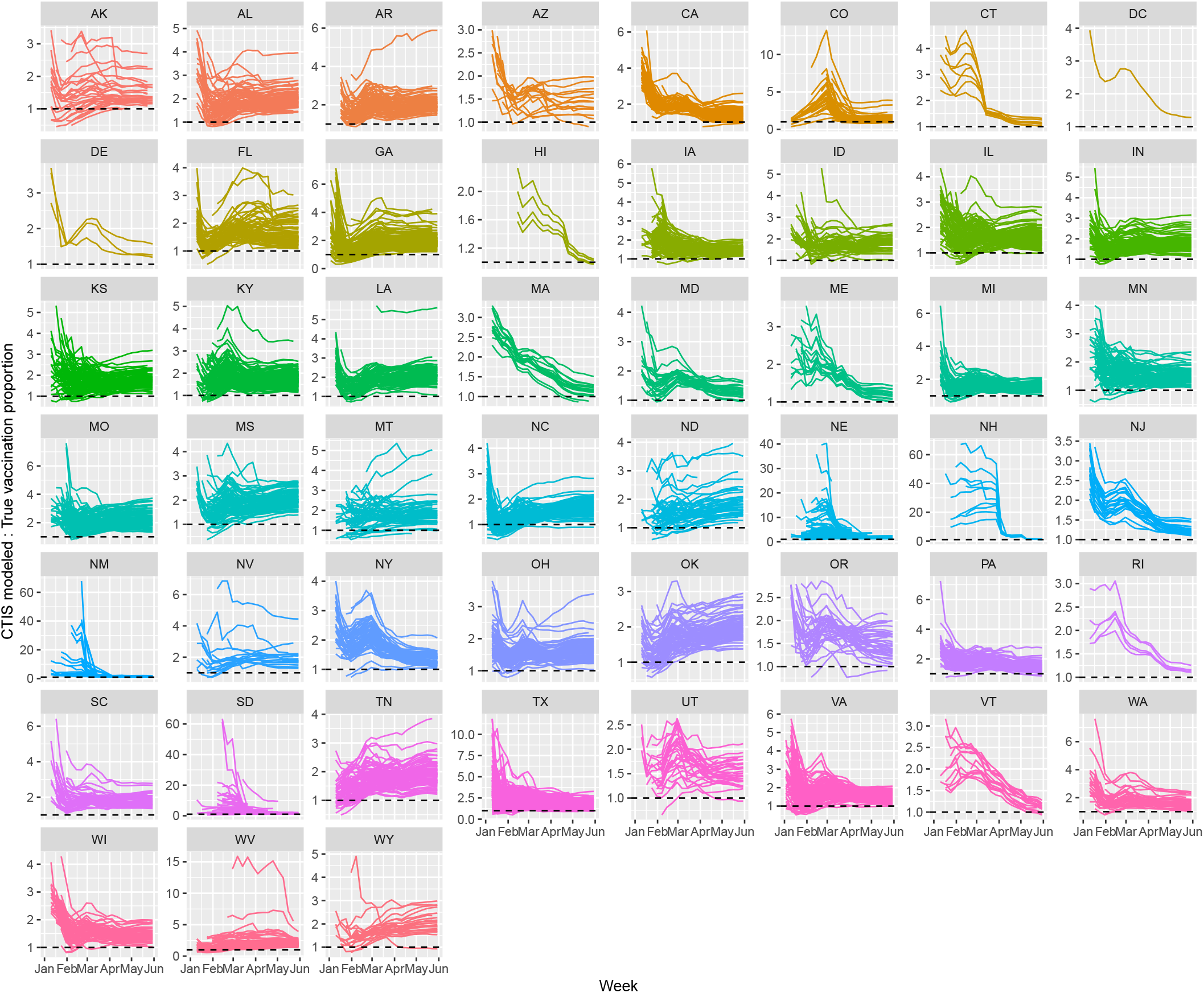
Motivation for bias correction using vaccination survey and ground-truth data. Ratio of modeled CTIS vaccination estimates to true vaccination proportions for each state over time (January 1 to June 30, 2021). Dashed line at a ratio of 1. CTIS survey estimates often substantially overestimate the fraction of vaccinated individuals.

**Figure S19.**
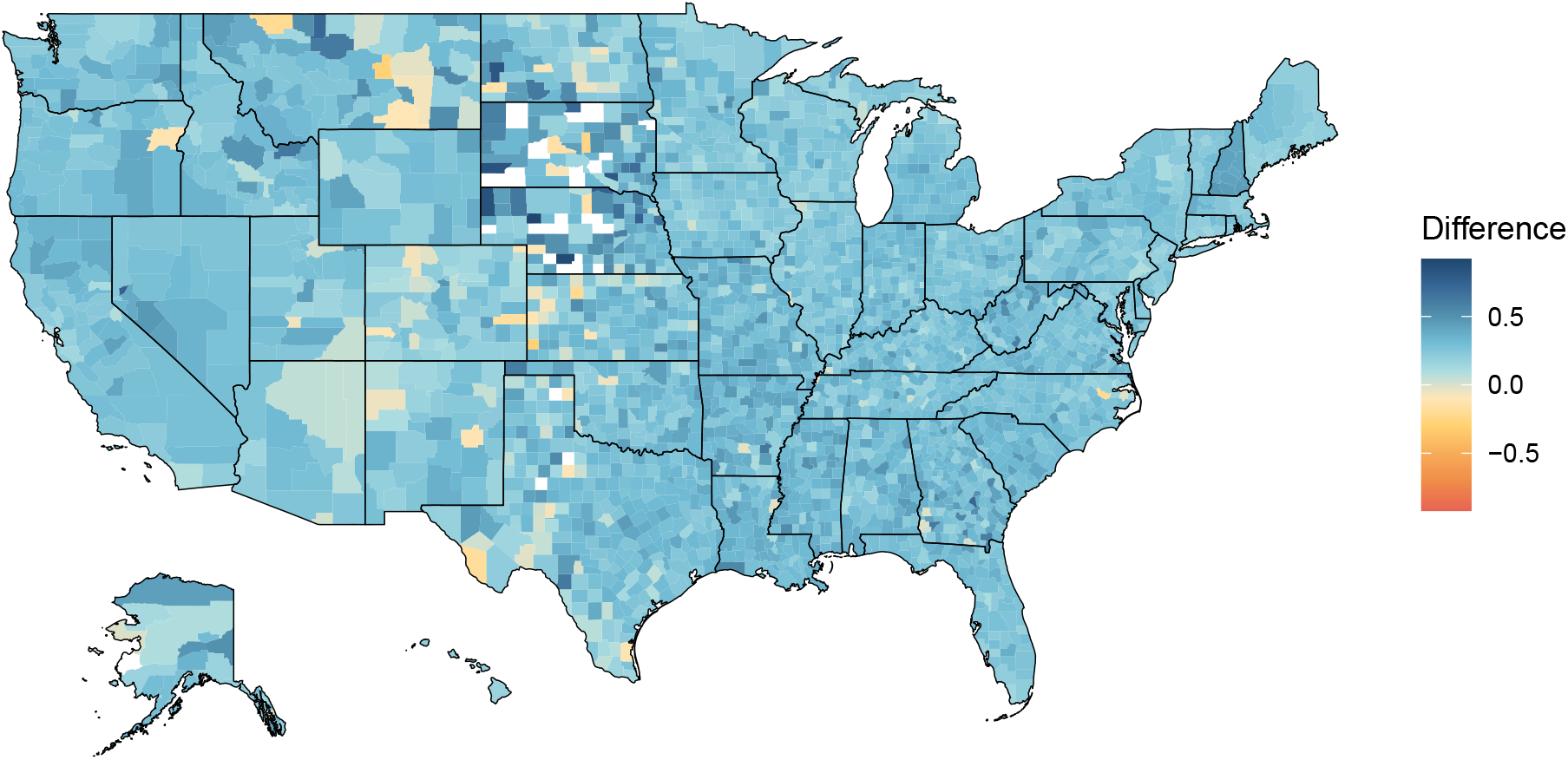
Map of differences between observed (raw) CTIS vaccination estimates and true COVID-19 vaccination coverage from the months of April and May. Note that these values are slightly different than the bias values used in our model which account for sample size issues in the CTIS vaccination estimates. Positive values indicate CTIS overestimated true vaccination coverage and negative values indicate underestimation of true coverage. There is some spatial heterogeneity in this bias.

**Figure S20.**
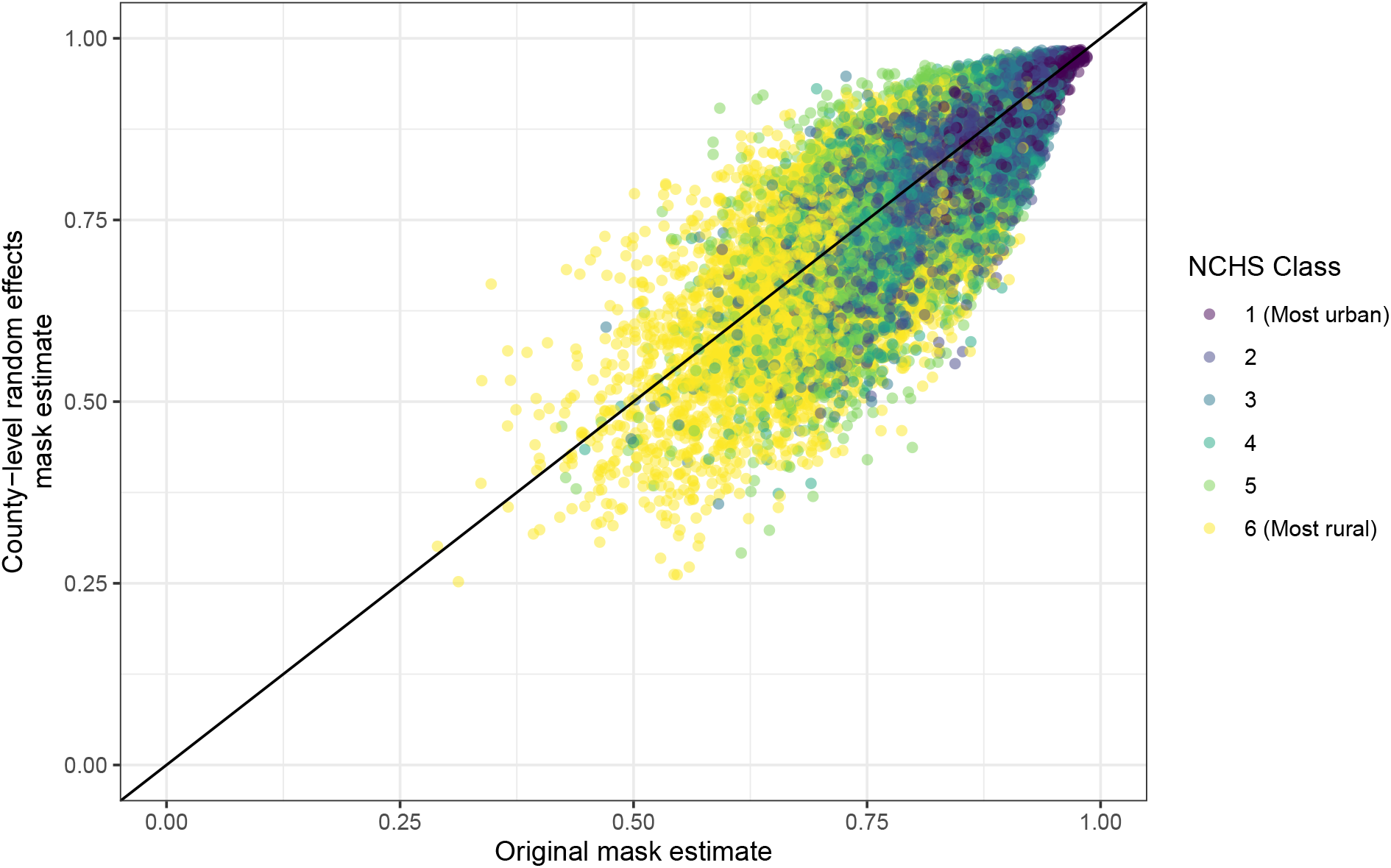
Estimates from model incorporating a county-level random effect (vertical axis) are, on average, similar to estimates from our original model without explicit spatial effects (horizontal axis). Both models use binomial regression on un-raked observations and do not incorporate the bias offset. Rural counties appear to show greater variability between the two models. Black line is y=x.

**Figure S21.**
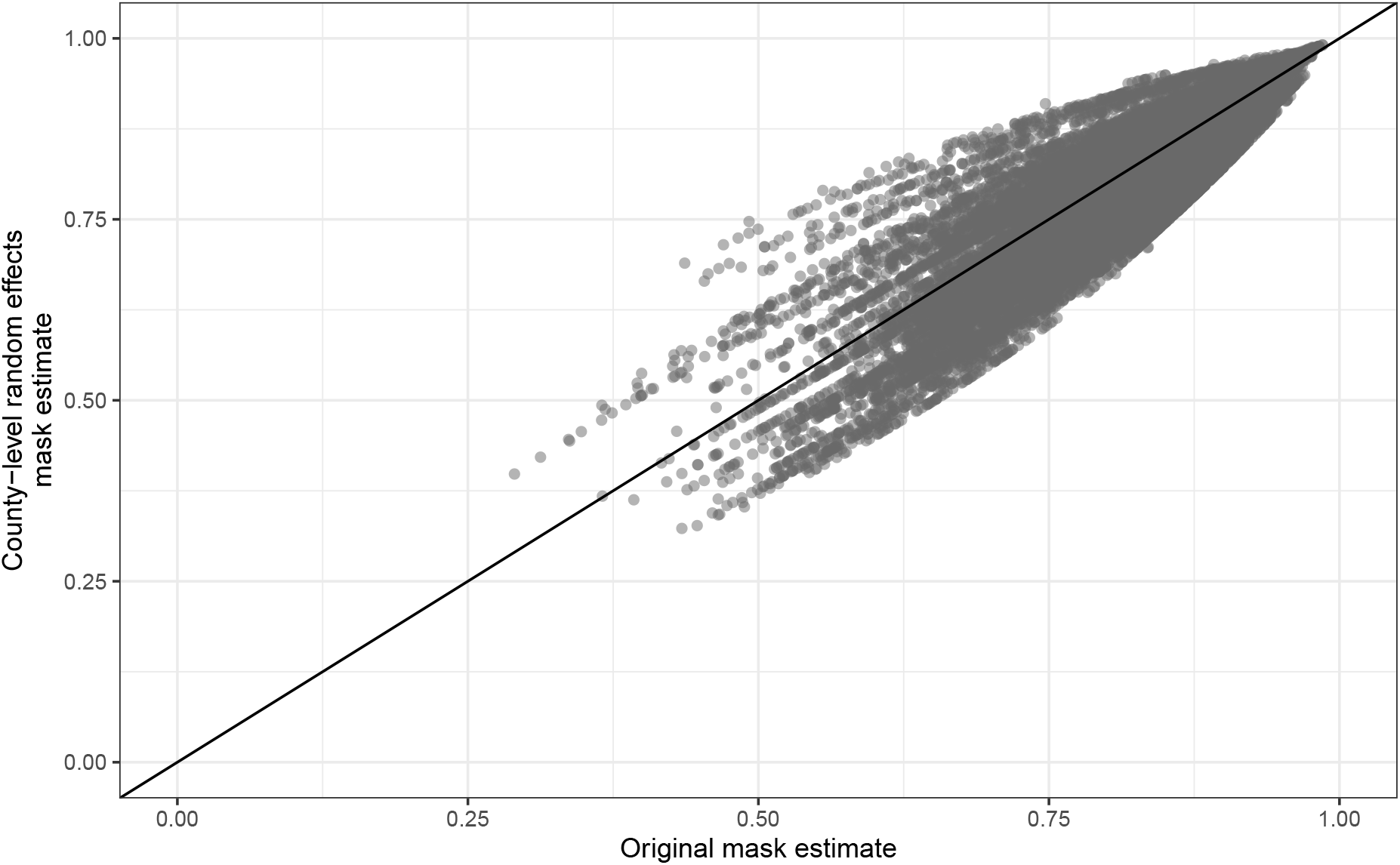
Estimates from model incorporating a state-level random effect (vertical axis) are, on average, similar to estimates from our original model without explicit spatial effects (horizontal axis). Both models use binomial regression on un-raked observations and do not incorporate the bias offset. The models with state-level random effects did not reliably converge and thus estimates should be interpreted with caution. Black line is y=x.

**Figure S22.**
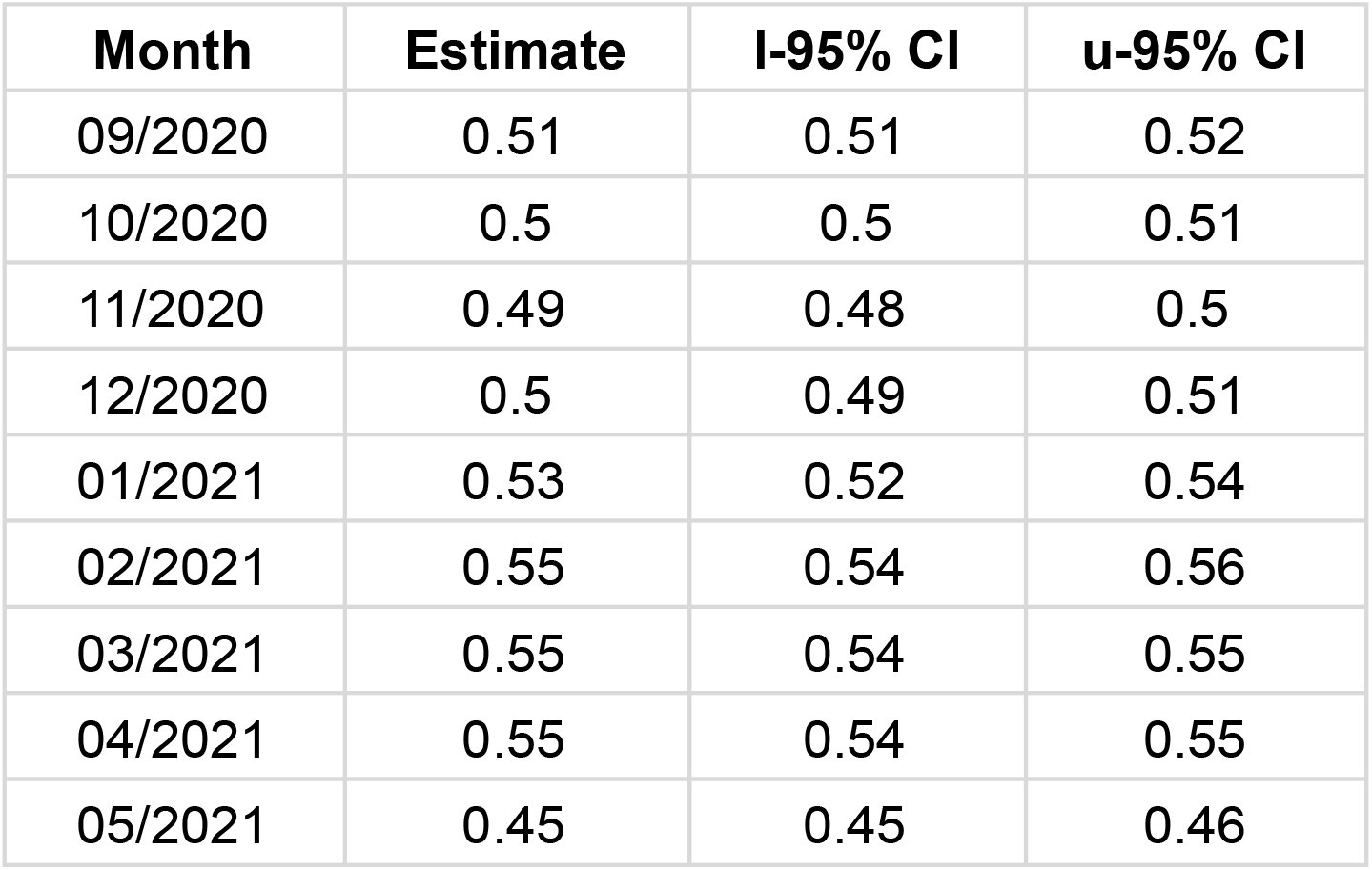
Coefficients and 95% credible intervals for z-score(*log*_10_(population density)) coefficient in the binomial regression model with raking and debiasing for each month. Coefficients are consistent over time.

**Figure S23.**
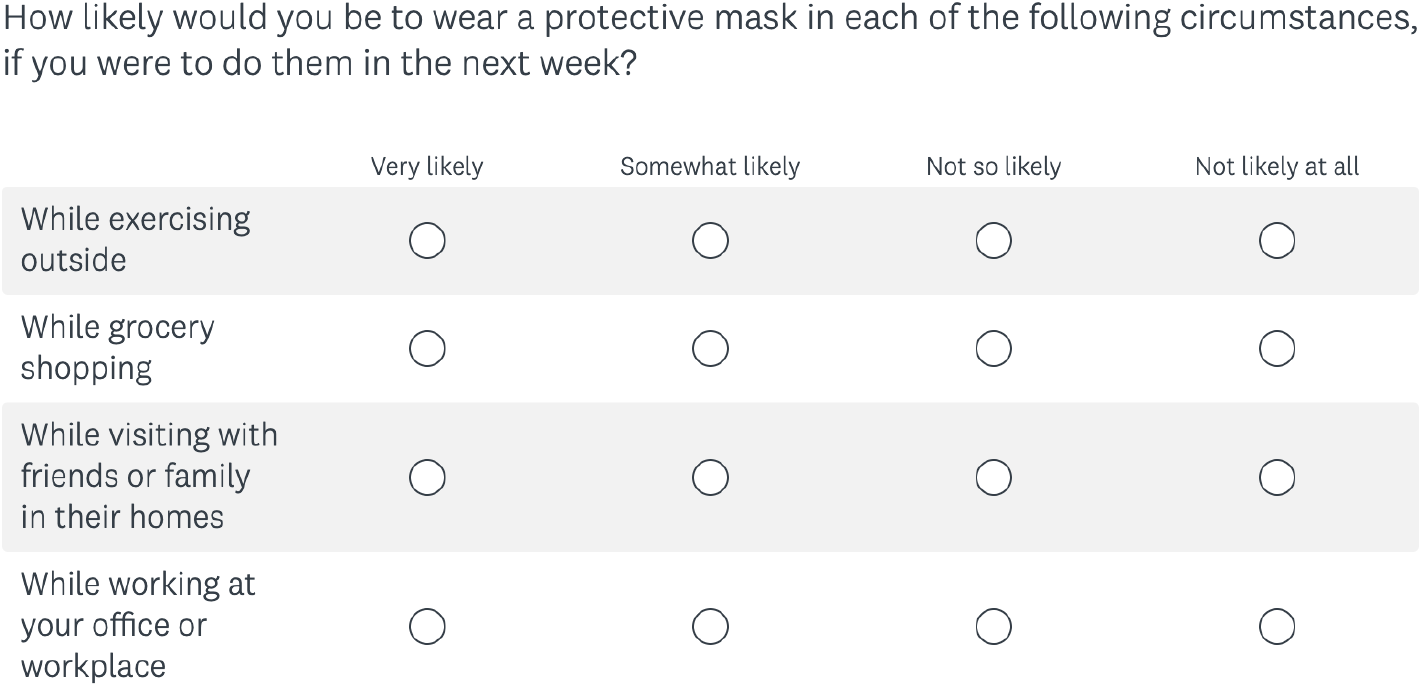
ONM question on masking behavior.

**Figure S24.**
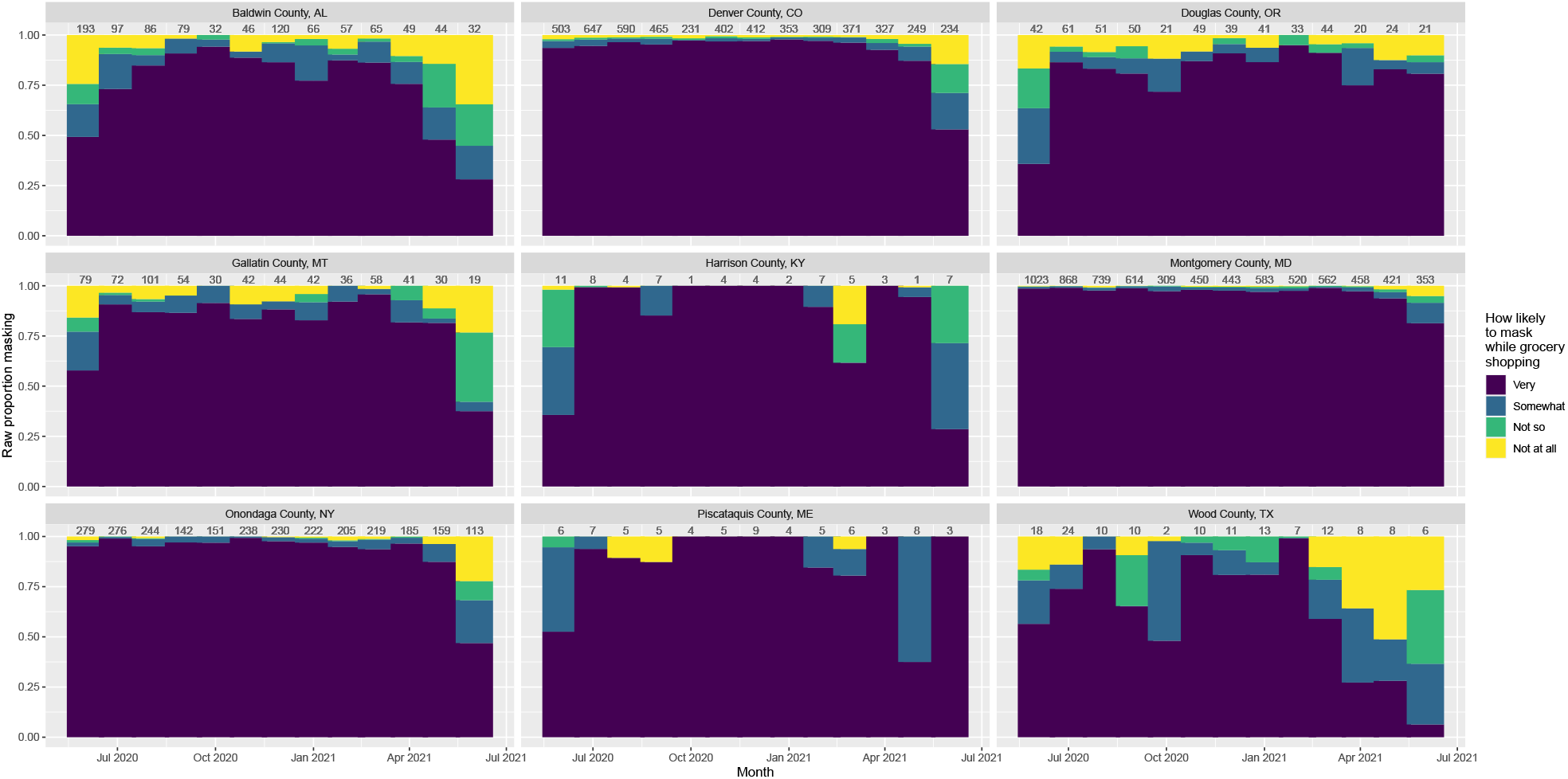
Raw proportions of each ONM response for a selection of counties spanning population densities across time. Number of responses in that county-month listed above each bar. NCHS urban-rural classifications are as follows: (1) Denver, CO; (2) Montgomery, MD; (3) Onondaga, NY; (4) Baldwin, AL; (5) Gallatin, MT; Douglas, OR; (6) Wood, TX; Piscataquis, ME; Harrison, KY.

**Figure S25.**
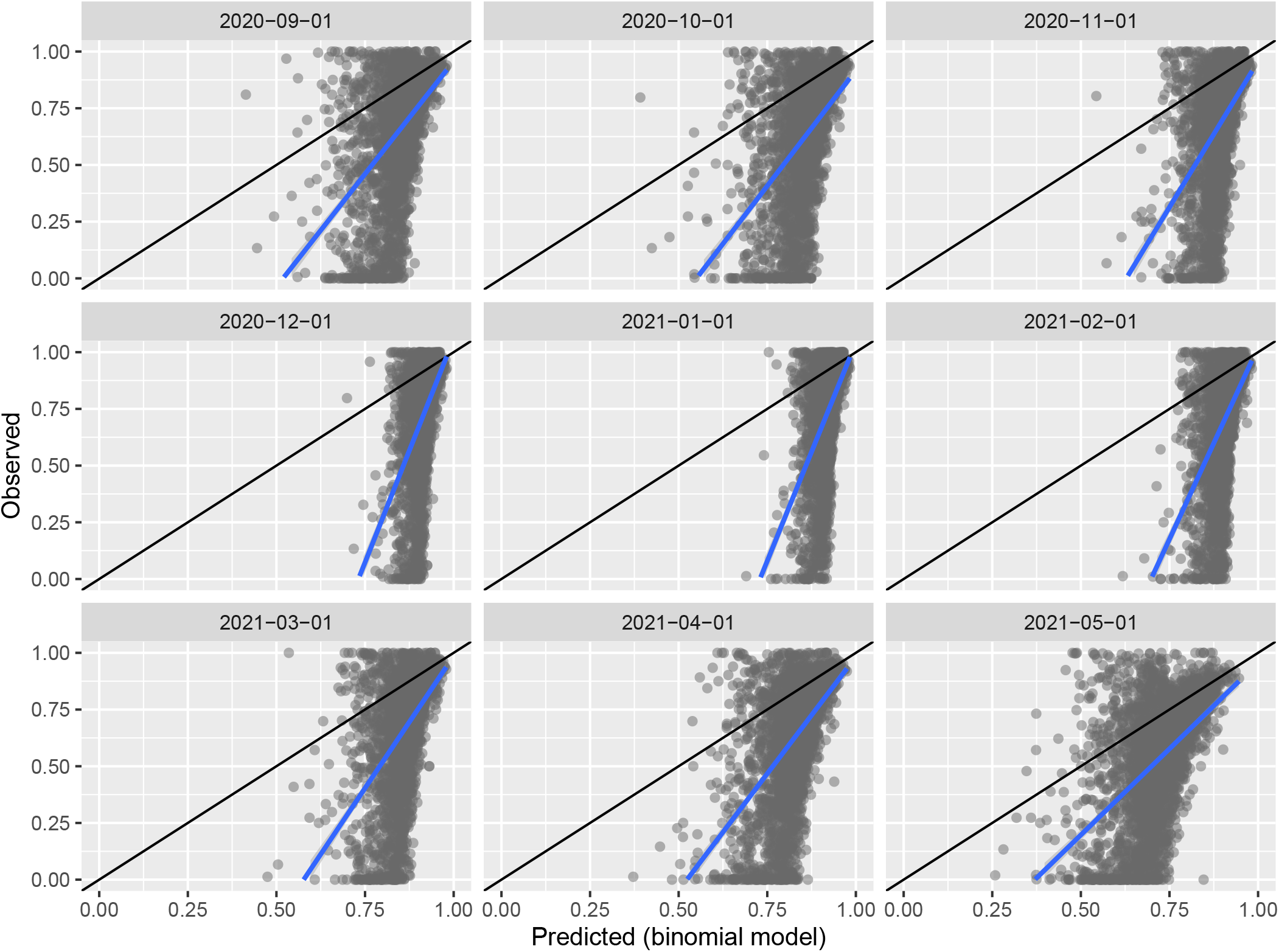
Observed versus predicted ONM masking estimates from binomial regression model. Black line shows y = x; blue line is a linear fit of the data. The linear fit is not close to the one-to-one line indicating poor model fit.

**Figure S26.**
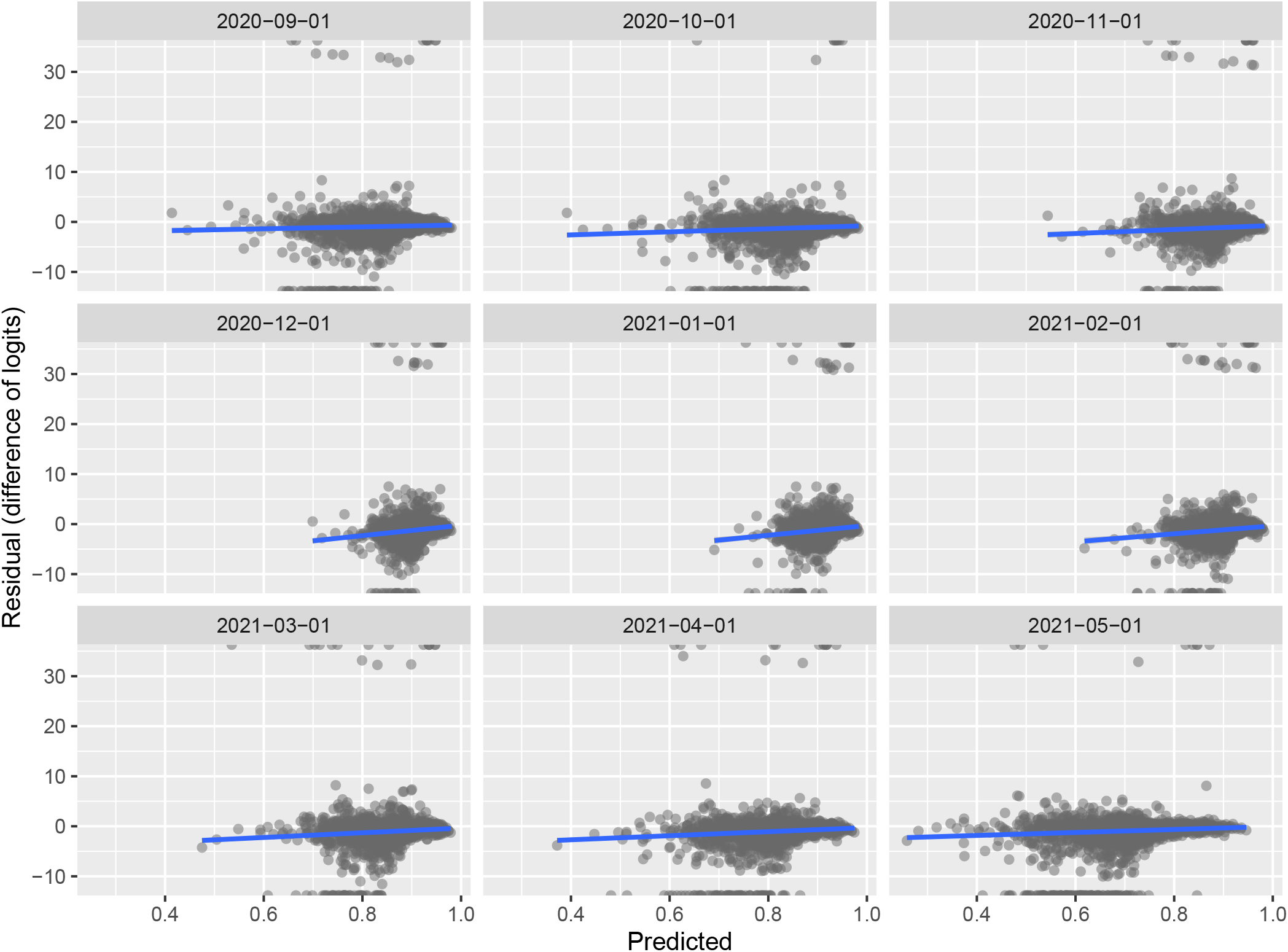
Observed versus residuals for ONM masking estimates. Blue line is a linear fit of the data. Residuals show weak association with predicted value, indicative of a mediocre model fit; there are also several outliers.

**Table S1.** Validation of model estimates from observational studies. Most studies that overlap with the survey period estimate the proportion of people wearing masks correctly within 10% of our debiased model estimates (green cells). Yellow cells denote literature estimates more than 10% higher than our estimates and red cells denote estimates more than 10% lower, with speculation on the reason for these differences provided in the similarly named column. Gray date cells indicate lack of overlap with our survey period.

| Date (MM/YYYY) | Location | Lit. estimate | Model debiased estimate | Difference | Model fips | Model date (MM/YYYY) | Speculation on any differences | DOI/PMID/URL |
| --- | --- | --- | --- | --- | --- | --- | --- | --- |
| 09/2020 | New Orleans, LA | 0.32 | 0.78 | -0.46 | 22071 | 09/2020 | Observations of Bourbon Street which may be a sample disproportionately composed of tourists, people going out during a pandemic who may be less likely to wear a mask | 10.1371/journal.pone.0261321 |
| 08/2020 | Philadelphia, PA | 0.43 | 0.85 | -0.42 | 42101 | 09/2020 | Time period different, mostly observed people outside at parks/streets where mask compliance may have been lower due to the outdoor nature | 10.1016/j.pmedr.2021.101449 |
| 06/2021 | Milwaukee, Waukesha, Ozaukee, Washington, and Sheboygan, WI | 0.26 | 0.61 | -0.35 | 55079, 55133, 55089, 55131, 55117 | 05/2021 | Time period is different, averaging across several counties in Wisconsin | 10.1101/2022.01.18.22269479 |
| 09/2020 | Times Square, NY | 0.65 | 0.91 | -0.26 | 36061 | 09/2020 | Sample may be disproportionately composed of tourists during a pandemic who may be less likely to wear a mask if choosing to travel during this time | 10.1371/journal.pone.0261321 |
| 05/2020 - 06/2020 | Wisconsin | 0.41 | 0.54 | -0.13 | Average all WI | 09/2020 | Time period different, observed grocery stores across 20 counties, model estimate is averaged over the entirety of Wisconsin | 10.1101/2020.06.09.20126946 |
| 03/2021 - 04/2021 | Indianapolis, IN | 0.74 | 0.85 | -0.11 | 18097 | 03/2021 - 04/2021 | Indoor basketball tournament (March Madness) may not be representative of typical people living in this area | 10.1001/jama.2021.14057 |
| 06/2020 - 07/2020 | Auburn-Opelika, AL | 0.55 | 0.65 | -0.10 | 1081 | 09/2020 | Time period different, single estimate not recorded so averaging across Fig 2 | 10.1108/S0895-993520220000029006 |
| 10/2020 | Philadelphia, PA | 0.77 | 0.86 | -0.09 | 42101 | 10/2020 | Average across all observations in outside and retail sites | https://public.tableau.com/app/profile/city-ofphiladelphia/viz/shared/MPS6SH482 |
| 06/2020 - 08/2020 | Portland, OR and Toronto, CAN | 0.68 | 0.77 | -0.09 | 41051 | 09/2020 | Time period different, higher levels of masking in Portland than Toronto but individual proportions not given | 10.1136/bmjopen-2021-049389 |
| 09/2020 | Philadelphia, PA | 0.76 | 0.85 | -0.09 | 42101 | 09/2020 | Average across all observations in outside and retail sites | https://public.tableau.com/app/profile/city-ofphiladelphia/viz/shared/MPS6SH482 |
| 12/2020 | Philadelphia, PA | 0.88 | 0.92 | -0.04 | 42101 | 12/2020 | Average across all observations in outside and retail sites | https://public.tableau.com/app/profile/city-ofphiladelphia/viz/shared/MPS6SH482 |
| 11/2020 | Philadelphia, PA | 0.87 | 0.89 | -0.02 | 42101 | 11/2020 | Average across all observations in outside and retail sites | https://public.tableau.com/app/profile/city-ofphiladelphia/viz/shared/MPS6SH482 |
| 09/2020 | Little Italy, NYC, NY | 0.9 | 0.91 | -0.01 | 36061 | 09/2020 |  | 10.1371/journal.pone.0261321 |
| 12/2020 | Louisville, KY | 0.86 | 0.87 | -0.01 | 21111 | 12/2020 |  | 10.1371/journal.pone.0248324 |
| 07/2020 | Honolulu, HI | 0.77 | 0.76 | 0.01 | 15003 | 09/2020 | Time period different | PMID: 32914093 |
| 09/2020 | Las Vegas, NV | 0.86 | 0.85 | 0.01 | 32003 | 09/2020 |  | 10.1371/journal.pone.0261321 |
| 11/2020 - 05/2021 | Marion County, IN | 0.86 | 0.84 | 0.02 | 18097 | 11/2020 - 05/2021 | Weekly data are available and are pretty consistent over time with a drop in last couple study weeks (May); anywhere between 80 - 90% usually | 10.1097/PHH.0000000000001467 |
| 11/2020 - 05/2021 | King County, WA | 0.872 | 0.81 | 0.06 | 53033 | 11/2020 - 05/2021 | Most observations in period from 12/2020 - 04/2021 | 10.1177/00333549221100795 |
| 05/2020 | Chittenden County, VT | 0.76 | 0.66 | 0.1 | 50007 | 09/2020 | Time period different | 10.1177/00333549211009496 |
| 09/2020 | Key West, FL | 0.66 | 0.56 | 0.1 | 12087 | 09/2020 |  | 10.1371/journal.pone.0261321 |
| 06/2020 - 07/2020 | Greensboro, NC | < 0.75 | 0.72 |  | 37081 | 09/2020 | Time period is different | 10.23954/osj.v7i2.3084 |
| 04/2021 - 05/2021 | Auburn-Opelika, AL | 0.74 | 0.64 | 0.10 | 1081 | 04/2021 - 05/2021 | Essential businesses, single estimate not recorded so averaging across Fig 3 | 10.1108/S0895-993520220000029006 |
| 08/2020 | Milwaukee, Waukesha, Ozaukee, Washington, and Sheboygan, WI | 0.96 | 0.7 | 0.26 | 55079, 55133, 55089, 55131, 55117 | 09/2020 | Time period is different, averaging across several counties in Wisconsin | 10.1371/journal.pone.0240785 |
| 09/2020 - 11/2020 | South and West | 0.77 |  |  |  | 10/2020 | Location too vague for comparison, university campuses with mask mandates, 0.86 wore masks, but only 0.77 properly, 0.92 indoors properly | 10.15585/mmwr.mm7006e1 |

